# COLI-NET: Fully Automated COVID-19 Lung and Infection Pneumonia Lesion Detection and Segmentation from Chest CT Images

**DOI:** 10.1101/2021.04.08.21255163

**Authors:** Isaac Shiri, Hossein Arabi, Yazdan Salimi, Amir Hossein Sanaat, Azadeh Akhavanalaf, Ghasem Hajianfar, Dariush Askari, Shakiba Moradi, Zahra Mansouri, Masoumeh Pakbin, Saleh Sandoughdaran, Hamid Abdollahi, Amir Reza Radmard, Kiara Rezaei-Kalantari, Mostafa Ghelich Oghli, Habib Zaidi

## Abstract

**Background:** We present a deep learning (DL)-based automated whole lung and COVID-19 pneumonia infectious lesions (COLI-Net) detection and segmentation from chest CT images.

**Methods:** We prepared 2358 (347’259, 2D slices) and 180 (17341, 2D slices) volumetric CT images along with their corresponding manual segmentation of lungs and lesions, respectively, in the framework of a multi-center/multi-scanner study. All images were cropped, resized and the intensity values clipped and normalized. A residual network (ResNet) with non-square Dice loss function built upon TensorFlow was employed. The accuracy of lung and COVID-19 lesions segmentation was evaluated on an external RT-PCR positive COVID-19 dataset (7’333, 2D slices) collected at five different centers. To evaluate the segmentation performance, we calculated different quantitative metrics, including radiomic features.

**Results:** The mean Dice coefficients were 0.98±0.011 (95% CI, 0.98-0.99) and 0.91±0.038 (95% CI, 0.90-0.91) for lung and lesions segmentation, respectively. The mean relative Hounsfield unit differences were 0.03±0.84% (95% CI, −0.12 – 0.18) and −0.18±3.4% (95% CI, −0.8 - 0.44) for the lung and lesions, respectively. The relative volume difference for lung and lesions were 0.38±1.2% (95% CI, 0.16-0.59) and 0.81±6.6% (95% CI, −0.39-2), respectively. Most radiomic features had a mean relative error less than 5% with the highest mean relative error achieved for the lung for the *Range* first-order feature (- 6.95%) and *least axis length* shape feature (8.68%) for lesions.

**Conclusion:** We set out to develop an automated deep learning-guided three-dimensional whole lung and infected regions segmentation in COVID-19 patients in order to develop fast, consistent, robust and human error immune framework for lung and pneumonia lesion detection and quantification.

## I. Introduction

The recent pandemic of severe acute respiratory syndrome coronavirus 2 (SARS-CoV-2) disease (COVID-19) is posing great health concerns globally (1, 2). The COVID-19 pandemic has resulted in loss of lives, health and economic issues (3). Although, a large number of trials have been conducted to produce vaccines and/or treat COVID-19, a specific vaccine or therapy is still lacking (4, 5). For the diagnosis of COVID-19, Reverse Transcription-Polymerase Chain Reaction (RT-PCR) is a high sensitive molecular test, but bears inherently a number of limitations (6, 7). Furthermore, previous studies have indicated that thoracic Computed Tomography (CT) is a fast and highly sensitive approach for COVID-19 detection and management (8, 9). In this regard, dedicated ultra low-dose CT scanning protocols were recently devised (10).

In connection with the use of CT in COVID-19 management, a wide range of qualitative and quantitative studies have been carried out for diagnostic, prognostic and longitudinal follow up of patients (11–14). In these studies, whole lungs or infectious lesions were analyzed and several patterns and features were found to have high diagnostic and prognostic value (13, 15–19). However, accurate segmentation of lungs and infectious pneumonia lesions remains challenging (20). Hence, segmentation is the main issue impacting the outcome of both qualitative and quantitative studies (12, 20, 21). Although several segmentation approaches including manual delineation, semi-automated (22) and fully automated (21) techniques have been applied to CT images for COVID-19 management, they are still facing serious challenges to produce robust and dependable outcomes.

In medical image segmentation, particularly whole 3D volumes definition and big data analysis, manual delineation requires experienced trained radiologists, is time consuming, labor-intensive, and suffers from inter- and intra-observer variability concerns (23, 24). Whole lung segmentation is a pivotal step for further analysis, including extraction of the percentage of infection, well aerated portion of the lung, and enabling radiomics and deep learning analysis of COVID-19 patients (15, 18). Conventional algorithms, including rule-based and atlas-based, performed relatively well on normal and mild disease chest CT, but might fail in COVID-19 patients lung segmentation because of different stage of disease with different levels of severity (20). Furthermore, developing a fully automatic tool for lung and pneumonia COVID-19 lesions is highly desired owing to rapid changes in appearance and manifestation at different stages of the disease (13, 20).

Artificial intelligence (AI) algorithms, particularly its two major subcategories, machine learning (ML) and deep learning (DL), have been widely used for medical image analysis (25–30) and more recently in the segmentation of lung and pneumonia infectious lesions from chest CT images of COVID-19 patients (16). These studies reported that AI improved the accuracy of lesion detection/segmentation and reduced the bias associated with conventional approaches. In a study by Zheng *et al.* (31), a weakly-supervised deep learning algorithm was applied to chest CT images for automatic COVID-19 detection. Fan *et al.* (32) presented a COVID-19 lung infection segmentation deep network (Inf-Net) based on semi-supervised learning. Furthermore, a number of deep learning algorithms, namely UNet, UNet++, V-Net, Attention-UNet, Gated-UNet and Dense-UNet were used for COVID-19 lesion detection and segmentation from chest CT images (33, 34).

CT images are commonly acquired on various scanner models using different imaging protocols, and as such, the resulting datasets are heterogeneous, which might lead to inaccuracy in the developed models. Training a robust and generalizable deep learning model requires a large clean annotated dataset (35). Owing to the relatively recent outbreak of COVID-19 pandemic, producing a large labeled COVID-19 image dataset is impractical. Transfer learning (TL) has received attention to address the lack of large datasets for the implementation of machine/deep learning-based algorithms (36, 37). Various TL-based strategies were used for transferring knowledge from different domains, including natural images to medical images to develop more robust and generalizable models (37).

In the present study, we developed a DL-based automated detection and segmentation of lung and COVID-19 pneumonia infectious lesions (COLI-Net) from chest CT images. In this work, large lung and COVID-19 lesions datasets and TL used to train a residual network (ResNet) for lung and pneumonia infectious lesions segmentation.

## II. Material and Methods

### Clinical studies

For lung and COVID-19 lesions segmentation, we prepared 2358 (347259, 2D slices) and 180 (17341, 2D slices) multi-centric and multi-vendor volumetric CT images with lung and COVID-19 lesion segmentations.

#### Lung datasets

For lung segmentation training, we used 2298 chest CT images (328205, 2D slices) with different pathologies from different centers, including 800 normal subjects without any lung abnormalities from Iran Center#1 (81347, 2D slices), 400 images of non-small cell lung carcinoma patients from Cancer Imaging Archive (TCIA) (38–40) (48568, 2D slices), 200 non-COVID-19 pneumonia (49465, 2D slices) and 898 (148825, 2D slices) RT-PCR positive COVID-19 patients from Iran Center#2. All images were manually segmented by experienced radiologists to delineate the lungs.

#### COVID-19 lesions datasets

For COVID-19 lesions segmentation training, we used 120 (9557, 2D slices) RT-PCR positive image datasets, including 90 (8338, 2D slices) datasets from 3 different centers in Iran (Centers#1, #2, #3) where the infectious lesions were manually segmented by experienced radiologists, in addition to 30 (1250, 2D slices) CT images from Russia (41).

#### Image preprocessing

Prior to network training, all images were cropped and resized to 296×216 matrix size. Image intensities were clipped between −1024 and 300 Hounsfield units and then normalized to a range [0 - 1.3].

#### Residual neural network

The residual network (ResNet) proposed by Li *et al.* (42, 43) built upon TensorFlow was used for lung and COVID-19 lesions segmentation. The ResNet is composed of 20 convolutional layers where different dilation factors were used for different levels of feature extraction (zero dilatation factor for low-level, two dilatation factors for medium-level, and four dilatation factors for high-level). Every two layers were linked together with residual connections (Figure 1). Non-square Dice was used as loss function. provides descriptive detail of ResNet.

**Figure 1.**
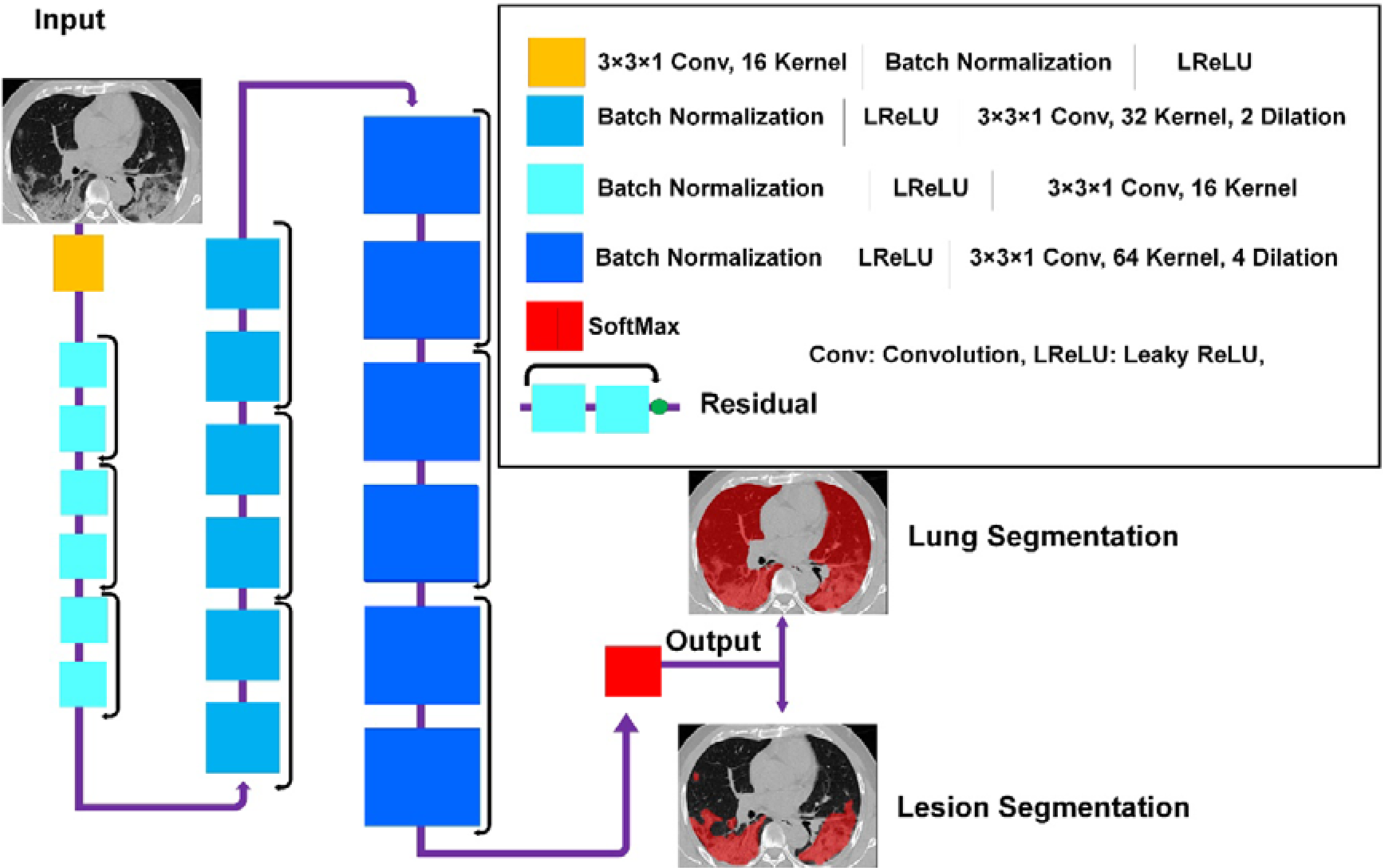
Architecture of the deep residual neural network (ResNet) along with details of the associated layers. Conv, convolutional kernel; LReLu, leaky rectified linear unit; SoftMax, Softmax function; Residual, residual connection.

#### Training and evaluation

Lung and COVID-19 lesions training was performed on 2D slices owing to the wide variability in slice thicknesses across the datasets from the different centers. For lung segmentation training, we used 2178 3D CT images (347259, 2D slices). For COVID-19 lesions segmentation, we used pretrained lung segmentation network as initial weights followed by fine-tuning for lesion segmentation of 120 3D CT images (9557, 2D slices). Body fine-tuning approaches were used for transfer learning where all pre-trained weights of lung segmentation were used as initial weights for lesion segmentation. The assessment of the quality of segmentations was performed independently on RT-PCR positive COVID-19 datasets from different centers, including 20 images (2214, 2D slices) from Center#1 (Iran1), 10 images (2552, 2D slices) from Center#2 (Iran2), 20 images (1250, 2D slices) from center#3 (Russia) (41), 10 images (939, 2D slices) from Center#4 (China) (44, 45) and 10 images (829, 2D slices) from center#5 (Italy^**1**^) (45). Overall, the evaluation was preformed on 7333 2D slices from different centers.

### Evaluation

To evaluate the performance of image segmentation, we calculated Dice similarity coefficient, Jaccard index, false negative, false positive, mean Hausdorff distance, and mean surface distance. In addition, different volume indices were exploited to quantify the portion of infection, including relative volume difference (%), relative volume difference of lesion/lung relative volume (%), absolute relative volume difference (%), absolute relative volume difference of lesion/lung relative volume (%). Hounsfield unit (mean) relative difference (%), and Hounsfield unit (mean) absolute relative difference (%) were calculated for lungs and COVID-19 lesions from different segmentations of CT images. Further details about the evaluated parameters are provided in the Supplemental Material. In addition, we evaluated the impact of the segmentation on 17 first-order and 10 shape radiomic features in both lungs and COVID-19 lesions. The list of radiomic features are presented in Supplemental Table 1.

## Results

Figures 2 and 3 compares visualy in 2D and 3D views for different external validation sets of lungs and lesions delineated manually by experienced radiologists and automatically by the deep learning model. Additional results from the external validation sets are provided in Supplemental Figures 1-13 (2D views) and 14-17 (3D views). Overall, there is good agreement between manual and predicted lung and infectious lesions segmentation in the different datasets. Despite the variability of the subjects among the different centers, COLI-Net performed consistently well in multi-centric and multi-scanner setting. What stands out from these results is that COLI-Net can detect and segment infectious regions (within lesion segmentation) while excluding arteries and tracheae in lung segmentation.

**Figure 2.**
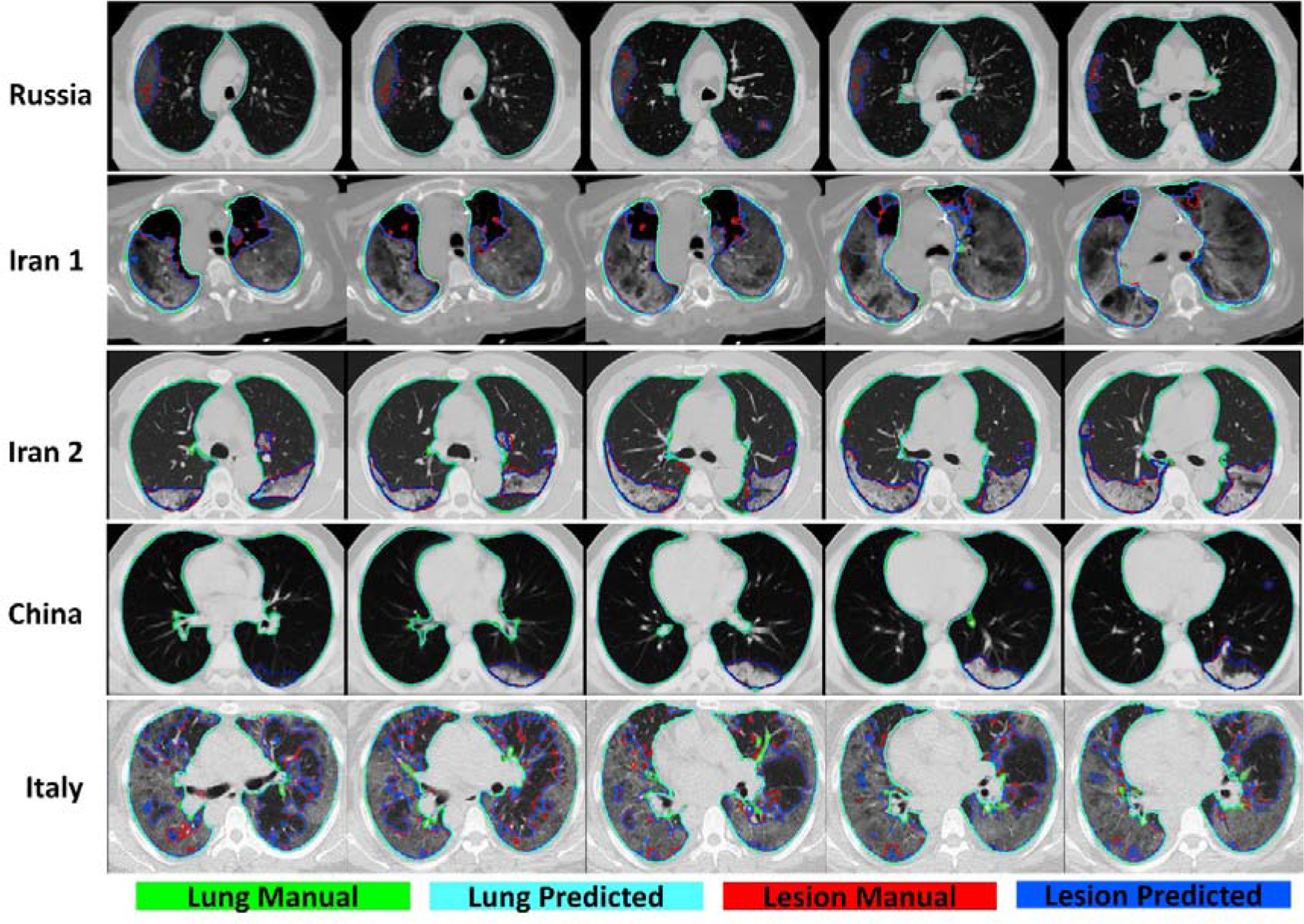
Representative manual and predicted segmentation (2D views) of lungs and COVID-19 lesions for 5 different cases from different datasets.

**Figure 3.**
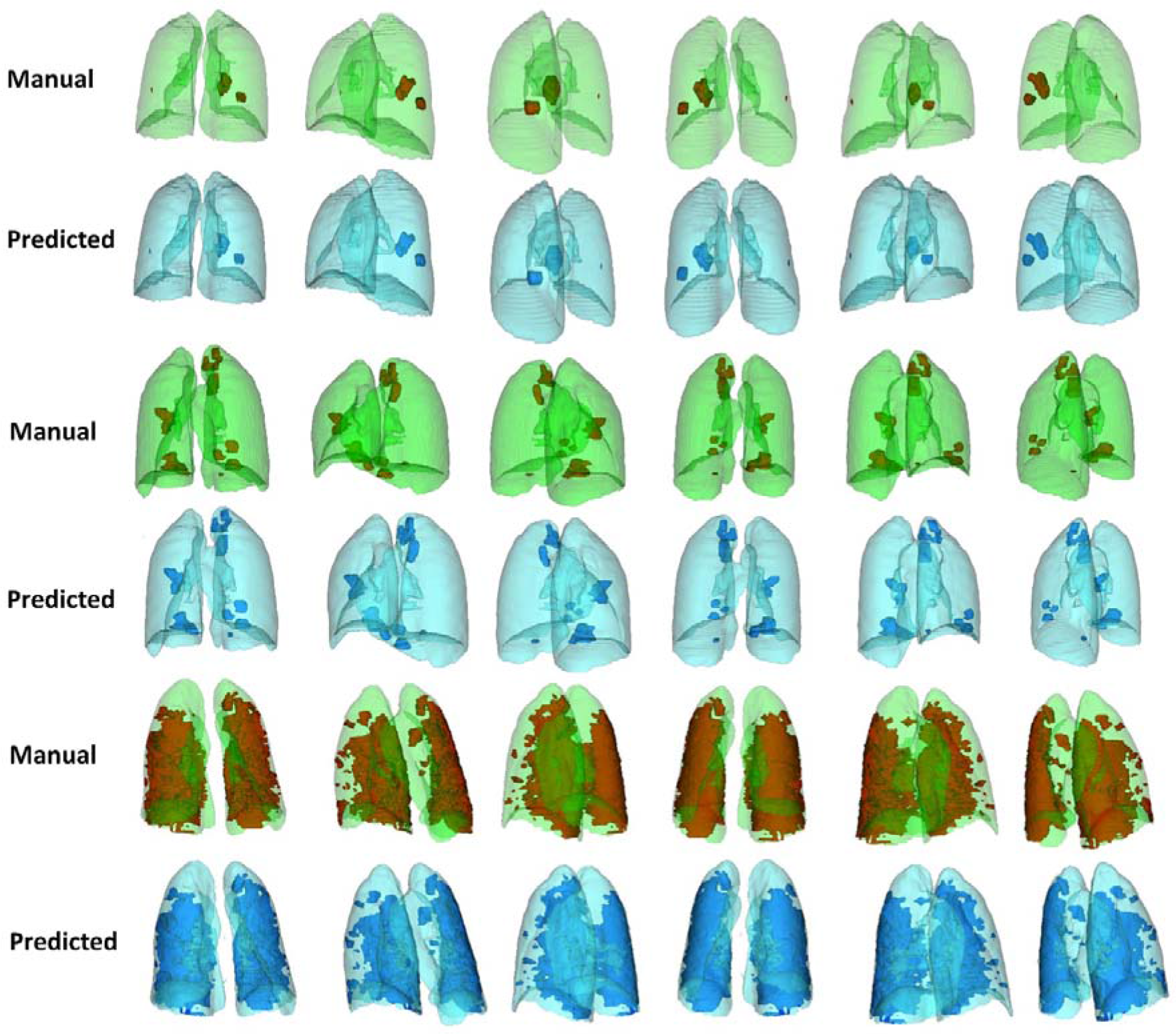
Representative manual and predicted segmentation (3D views) of lungs and COVID-19 lesions for 3 different cases from different datasets.

Table 1 summarizes segmentation quantification metrics for lungs and COVID-19 lesions. It can be seen that the mean Dice coefficients were 0.98±0.011 (95% CI, 0.98-0.99) and 0.91±0.038 (95% CI, 0.90-0.91) for lung and lesions segmentation, respectively. The mean Jaccard index was 0.97±0.022 (95% CI, 0.97-0.97) and 0.83±0.062 (95% CI, 0.82-0.84) for lung and COVID-19 lesions segmentation, respectively. Supplemental Tables 2-7 summarize lung and lesion segmentation quantification metrics for different external validation sets.

**Table 1.** Descriptive statistics of quantitative metrics for lung and COVID-19 lesions in the different datasets.

|  | **Metric** | **Min** | **Max** | **Mean ± SD** | **95% CI** |
| --- | --- | --- | --- | --- | --- |
| **Lung** | Dice | 0.92 | 0.99 | $0.98 \pm 0.011$ | 0.98 - 0.99 |
| | Jaccard | 0.86 | 0.99 | $0.97 \pm 0.022$ | 0.97 - 0.97 |
| | False Negative | 0.003 | 0.086 | $0.013 \pm 0.011$ | 0.011 - 0.015 |
| | False Positive | 0.002 | 0.073 | $0.017 \pm 0.014$ | 0.014 - 0.019 |
| | Average Hausdorff Distance | 0.005 | 0.14 | $0.022 \pm 0.026$ | 0.018 - 0.027 |
| | Mean Surface Distance | 0.005 | 0.17 | $0.026 \pm 0.028$ | 0.021 - 0.031 |
| **Lesions** | Dice | 0.8 | 0.98 | $0.91 \pm 0.038$ | 0.9 - 0.91 |
| | Jaccard | 0.66 | 0.96 | $0.83 \pm 0.062$ | 0.82 - 0.84 |
| | False Negative | 0.015 | 0.23 | $0.086 \pm 0.044$ | 0.078 - 0.094 |
| | False Positive | 0.024 | 0.32 | $0.098 \pm 0.055$ | 0.088 - 0.11 |
| | Average Hausdorff Distance | 0.043 | 5.6 | $0.42 \pm 0.73$ | 0.29 - 0.55 |
| | Mean Surface Distance | 0.046 | 6.1 | $0.45 \pm 0.79$ | 0.31 - 0.59 |

**Table 2.** Descriptive statistics of volume index for lung and COVID-19 lesions in the different datasets.

|  | **Metric** | **Min** | **Max** | **Mean ± SD** | **95% CI** |
| --- | --- | --- | --- | --- | --- |
| **Lung** | Relative Mean HU Diff (%) | -4.2 | 3.9 | $0.03 \pm 0.84$ | -0.12 - 0.18 |
| | Absolute Relative Mean HU Diff (%) | 0.006 | 4.2 | $0.52 \pm 0.66$ | 0.4 - 0.64 |
| | Relative Volume Diff (%) | -3.1 | 6.4 | $0.38 \pm 1.2$ | 0.16 - 0.59 |
| | Absolute Relative Volume Diff (%) | 0.004 | 6.4 | $0.89 \pm 0.88$ | 0.73 - 1 |
| **Lesions** | Relative Mean HU Diff (%) | -9.8 | 10 | $-0.18 \pm 3.4$ | -0.8 - 0.44 |
| | Absolute Relative Mean HU Diff (%) | 0.026 | 10 | $2.4 \pm 2.5$ | 1.9 - 2.8 |
| | Relative Volume Diff (%) | -14 | 21 | $0.81 \pm 6.6$ | -0.39 - 2 |
| | Absolute Relative Volume Diff (%) | 0.018 | 21 | $4.8 \pm 4.6$ | 4 - 5.6 |

Table 2 summarizes the impact of lung and lesions segmentations on mean Hounsfield unit and volume calculation. Mean relative HU differences (%) of 0.03±0.84 (95% CI, −0.12 – 0.18) and −0.18 ± 3.4 (95% CI, −0.8 - 0.44) were achieved for lungs and lesions, respectively. The relative volume difference for the lung was 0.38±1.2 (95% CI, 0.16-0.59) whereas it was 0.81±6.6 (95% CI, −0.39-2) for lesions. The results obtained from the mean Hounsfield unit and volume calculation for lung and infectious lesions for the different external validation sets are presented in Supplemental Tables 8-11.

Figures 4 and 5 depict the Dice similarity index, Jaccard, mean Hounsfield unit, and volume difference box plots for lung and lesions segmentation, respectively. Supplemental Figures 18 and 19 show box plots of Hounsfield unit absolute relative difference (%), absolute relative volume difference (%), false negative, false positive, average Hausdorff distance, and mean surface distance for lung and lesions.

**Figure 4.**
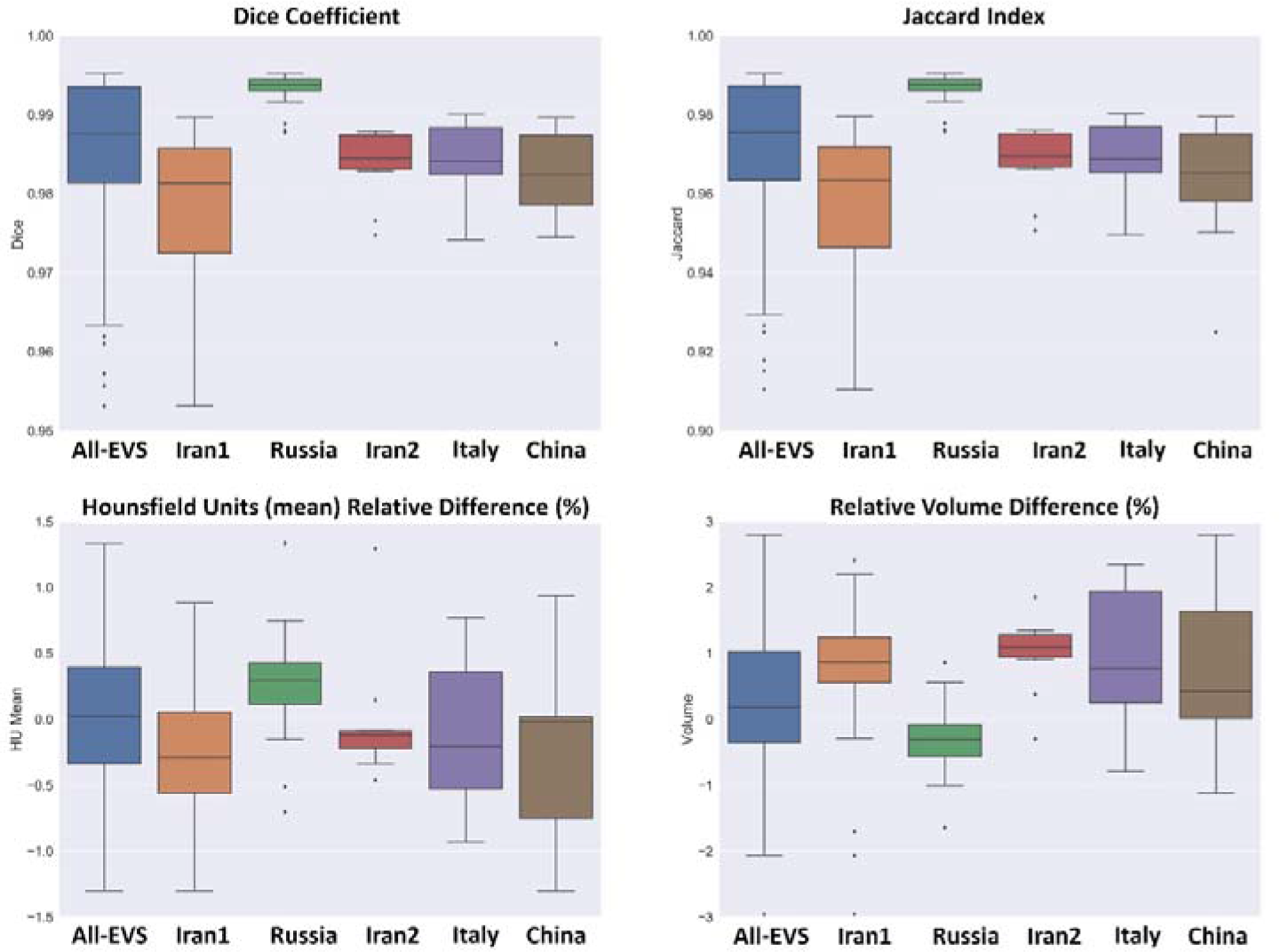
Box plots comparing various quantitative imaging metrics for lung segmentation, including Dice coefficient, Jaccard index, Hounsfield units (mean) relative difference (%) and relative volume difference (%).

**Figure 5.**
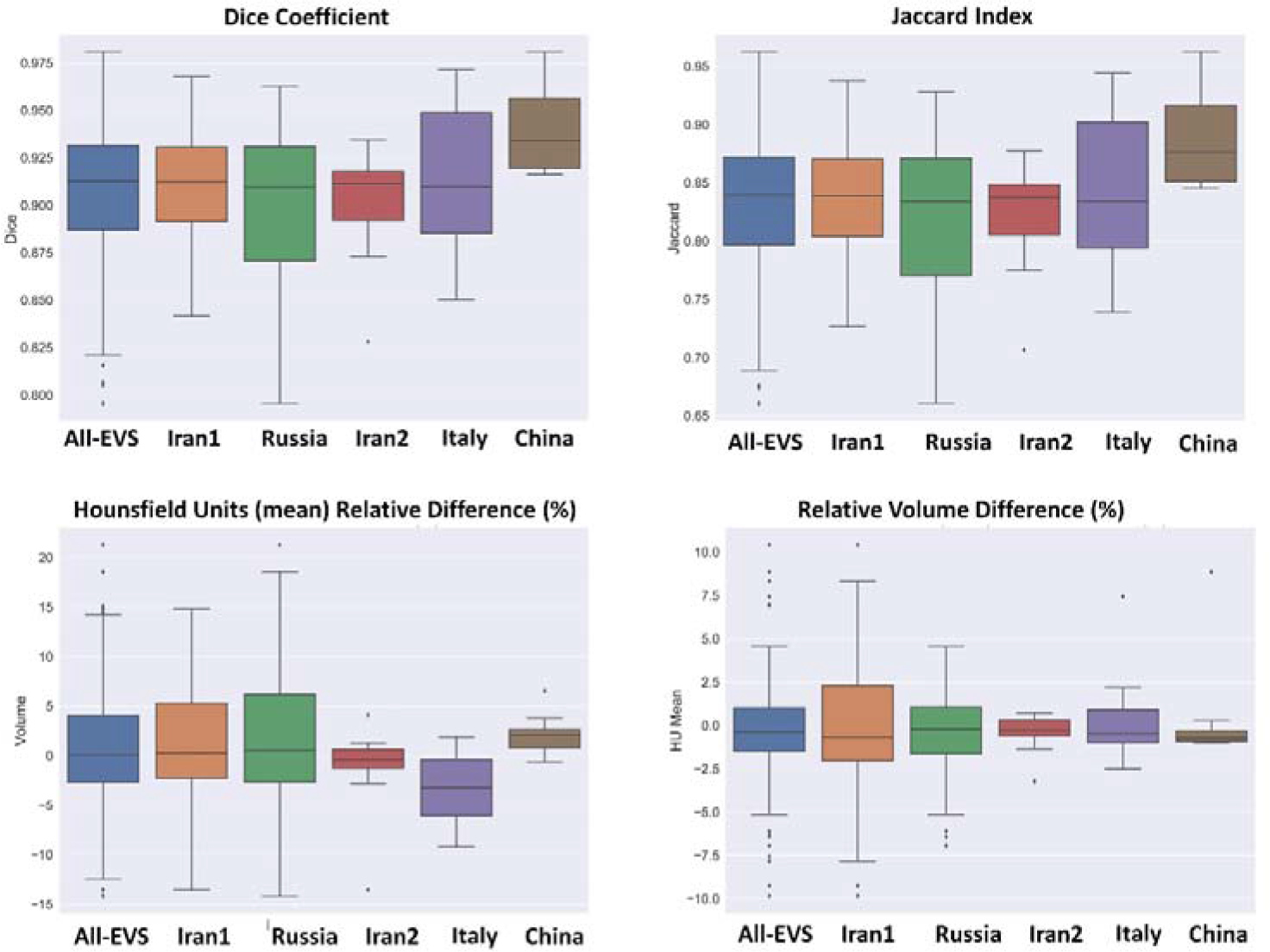
Box plots comparing various quantitative imaging metrics for COVID-19 lesions segmentation, including Dice coefficient, Jaccard index, Hounsfield units (mean) relative difference (%) and relative volume difference (%).

Descriptive statistics of relative volume (lesion/lung) indices are presented in Table 3. A relative error of 0.22±6.3 (95% CI, −0.95-1.4) and absolute relative error of 4.7±4.2 (95% CI, 3.9-5.5) were achieved for relative volume (lesion/lung). Supplemental Tables 12 and 13 summarize the results obtained for the relative volume (lesion/lung) index for different external validation sets. Figure 6 decpits boxplot of manual and predicted relative volume lesion/lung differences (%) and absolute/relative error of lesion/lung relative volumeerrors (%) for different external validation sets.

**Figure 6.**
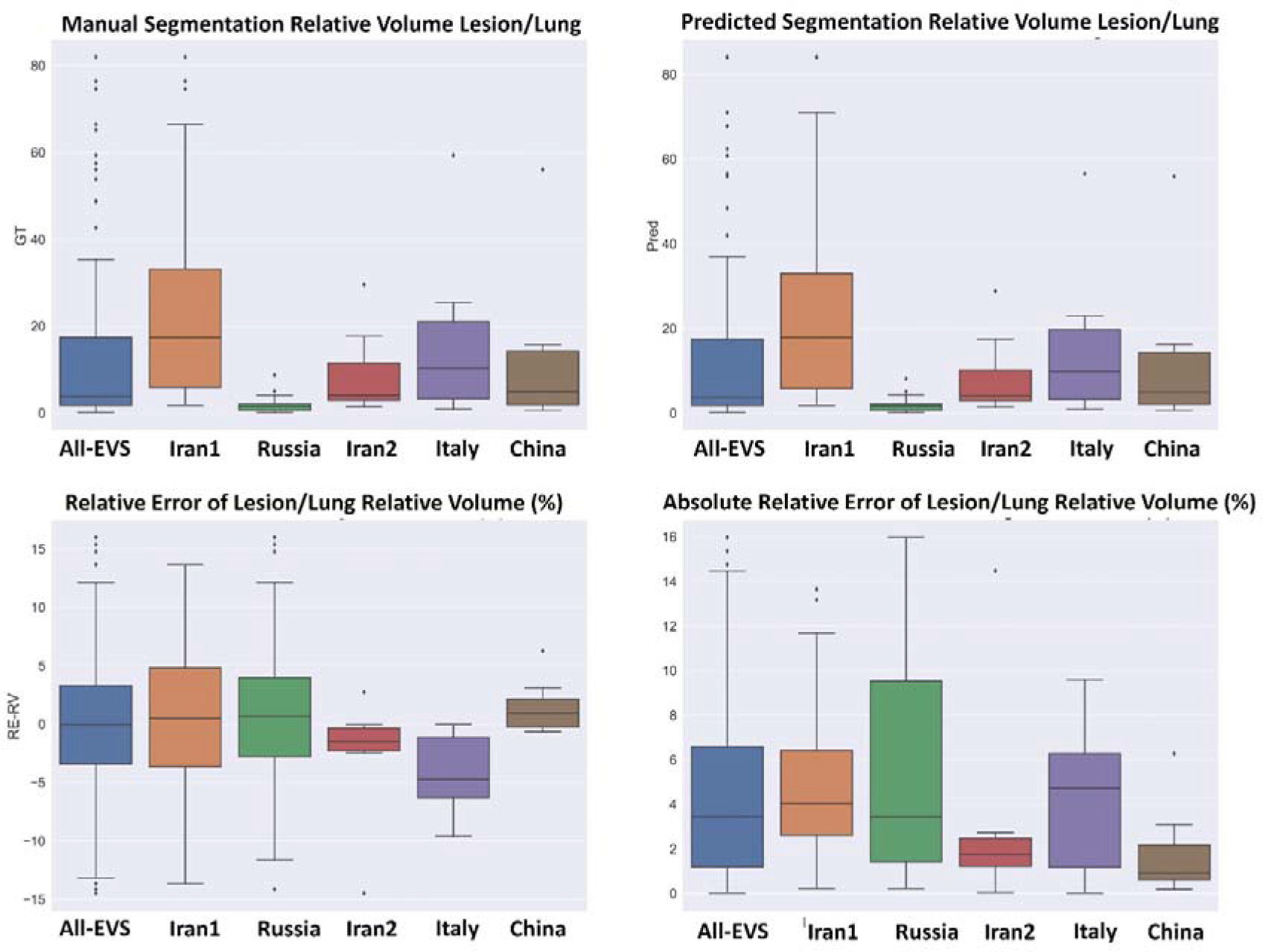
Box plots comparing various quantitative imaging metrics for relative volume, including manual segmentation relative volume lesion/lung, predicted segmentation relative volume lesion/lung, relative error of lesion/lung relative volume (%) and absolute relative error of lesion/lung relative volume (%).

**Table 3.** Descriptive statistics of relative volume index.

| Metric | Min | Max | Mean ± SD | 95% CI |
| --- | --- | --- | --- | --- |
| Manual segmentation relative volume (Lesion/Lung) | 0.001 | 0.82 | $0.13 \pm 0.19$ | 0.095 - 0.16 |
| Predicted segmentation relative volume (Lesion/Lung) | 0.001 | 0.84 | $0.13 \pm 0.19$ | 0.094 - 0.16 |
| RE Volume Diff Lesion/Lesion (%) | -14 | 16 | $0.22 \pm 6.3$ | -0.95 - 1.4 |
| ARE Volume Diff Lesion/Lesion (%) | 0.004 | 16 | $4.7 \pm 4.2$ | 3.9 - 5.5 |

Figure 7 presents heatmap of the mean relative error of first-order and histogram shape radiomic features in the lung and lesions for different validation sets. Most radiomic features exhibited a mean relative error less than 5% with the highest mean relative error for the lung being −6.95% for Range first-order feature and least axis length shape feature (8.68%) in lesions. The heatmap of the mean absolute relative error is depicted in Supplemental Figure 20.

**Figure 7.**
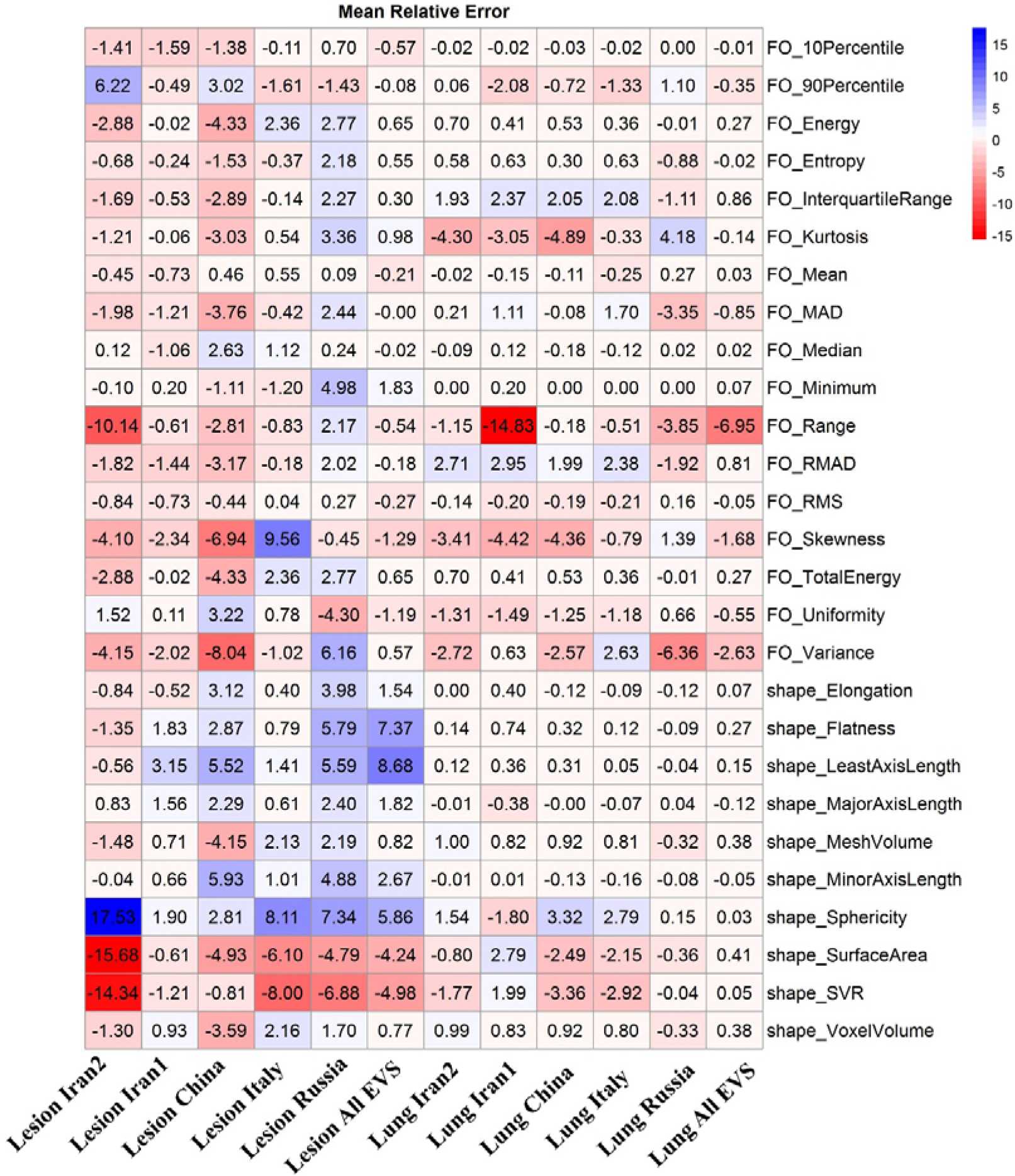
Mean relative error of different first-order and shape radiomic features for different datasets in lung and infection regions.

## Discussion

Chest CT imaging has emerged as a complementary tool for COVID-19 early diagnosis and longitudinal follow up (8). However, a number of challenges still need to be addressed for the accurate diagnosis of COVID-19 and its differentiation from other lung diseases, such as viral and bacterial pneumonia and other respiratory diseases (18). In this regard, several AI-based solutions exhibiting different levels of accuracy and robustness were proposed and evaluated (15, 18).

Another challenging problem that arises in the domain of quantitative analysis of CT images in clinical practice is lung and pneumonia infectious lesions segmentation (20). At the outset, different complex manifestations (appearance, size, location, boundaries and contrast) of infectious lesions, including consolidation, reticulation, and Ground-Glass Opacity (GGO) at different stages of the disease (longitudinal changes in the same patients) have been observed (13). Furthermore, providing ground truth segmentation for infectious lesion segmentation is challenging owing to inter-/intra-observer variability, noisy annotations, and the long processing time (20).

Previously developed atlas (46), rule (47), and hybrid (atlas and rule) (48) based algorithms for lung segmentation have shown acceptable performance on normal lungs and in the presence of mild pathogens (low density), such as emphysema (49). However, they presented limited performance in severe conditions (high density), including pleural effusion, atelectasis, consolidation, fibrosis, and pneumonia (50). Recent developments in the field of machine learning have led to a renewed interest in automatic lung segmentation. However, most seminal works in this area used a limited training dataset, predominantly containing normal cases or focusing on one class of pathogeneses, which could impact generalizability for unseen/non-diagnoised test datasets (51). In the present study, we applied deep learning algorithms and transfer learning on CT images obtained from different imaging centers to detect and segment the whole lung and pneumonia infected regions in COVID-19 patients.

A number of works attempted to develop automatic segmentation algorithms for lung and infectious lesions in COVID-19 CT images. Hofmanninger *et al.* (50) developed models for lung segmentation and reported a dice of 0.98 ± 0.01 for different pathological states (atelectasis, fibrosis, mass, pneumothorax, and trauma). They concluded that diversity in the training dataset is more important than deep learning algorithms. Müller *et al.* (52) implemented a 3D U-Net using data augmentation for generating image patches during training for lung and lesion segmentation on 20 annotated CT volumes. They achieved Dice coefficients of 0.950 and 0.761 for lung and lesions, respectively. A modified 3D U-Net (feature variation and progressive atrous spatial pyramid pooling blocks) proposed by Yan *et al.* (33) was developed for lung and infectious lesion segmentation on 861 patients, reporting a Dice similarity index of 0.987 for lung and 0.726 for lesions segmentation. Moreover, comparisons were performed with a fully dense Fully Convolutional Network (FCN) (lung: 0.865, lesions: 0.659) (53), U-Net (lung: 0.987, lesions: 0.688) (54), V-Net (lung: 0.983, lesions: 0.625) (55), and U-Net++ (lung: 0.986, lesions: 0.681) (56). The mean Dice for lung and lesions segmentation for different external validation sets used in our work are 0.98±0.011 and 0.91±0.038, respectively.

Chen *et al.* (57) used the residual attention U-Net for multi-class segmentation of CT images, achieving a Dice of 0.94 for infectious lesions segmentation. Zhou *et al.* (34) used a modified U-net network through spatial and channel attention mechanisms along with focal Tversky loss in the training process for improving small lesions segmentation. The results were evaluated on 427 slices achieving a Dice of 0.83. Elharrouss *et al.* (58) adopted an encoder-decoder for infectious lesions segmentation and used 20 clinical studies from the Italian Society of Medical and Interventional Radiology to report a Dice of 0.786. They compared the results with U-Net (Dice: 0.439) (59), Attention-UNet (Dice: 0.583) (60), Gated-UNet (Dice: 0.623) (61), Dense-UNet (Dice: 0.515) (62), U-Net++ (Dice: 0.422) (56) and Inf-Net (Dice: 0.739) (32). Wang *et al.* (63) proposed a robust algorithm for COVID-19 infectious lesions segmentation from CT images (COPLE-Net) designed to learn from noisy labeled data. The algorithm relies on noise-robust Dice loss and mean absolute error loss for generalized Dice loss for robust segmentation of noisy datasets and a modified version of U-Net to better handle infectious lesion segmentation with various manifestations and scales. The best results achieved by COPLE-Net were 0.807±0.099 and 0.160±0.171% as Dice coefficient and relative volume error (RVE (in %)) respectively. Wang *et al.* (63) evaluated different deep learning algorithms, including modified 3D U-Net (3D New-Net U-Net, Dice: 0.704±0.187, RVE: 25.41±24.73%) (64), modified 2D U-Net (2D New-Net U-Net, Dice: 0.791±0.129, RVE: 18.37±17.43%) (64), spatial attention gate U-Net (Attention U-Net, Dice: 0.772±0.123, RVE: 19.77±18.41%) (60), spatial and channel ‘squeeze and excitation’ blocks with U-net (ScSE U-Net, dice: 0.780±0.125, RVE: 18.85±16.69%) (65), and light-weight power efficient and general purpose CNN (ESPNetv2, Dice: 0.698±0.148, RVE: 23.69±20.26%) (66). Our proposed COLI-Net showed good performance compared to previous studies with a a Dice of 0.91±0.038 (95% CI: 0.90 - 0.91) and RVE of 0.38 ± 1.2% (95% CI: 0.16 - 0.59) for pneumonia infectious lesions.

A large labeled dataset is required to build a robust and generalizable model and avoid overfitting. Previous studies attempted to transfer the knowledge from natural to medical imaging domain, leading to improved accuracy by addressing the issue of limited datasets (36, 37). Transfer learning was recently applied for the detection and classification of COVID-19 using chest x-ray and CT images (67, 68). More recently, Wang *et al.* (69) applied four transfer learning methods on COVID-19 CT images for the segmentation of infectious lesions using 3D U-Net. The information was transformed from cancer and pleural effusion data to COVID-19 lesion segmentation, the Dice coefficient increased from 0.673±0.22 to 0.703±0.20 after transfer learning (69). They concluded that the transferability of non-COVID-19 data improved the quality of COVID-19 lesion segmentation to build a robust segmentation model. In our study, we exploited transfer learning from a large multicentric lung labeled dataset with various pathologies to overcome the shortcomings of infectious lesion segmentation.

Li *et al.* (70) used thick-section chest CT images of 531 COVID-19 patients for automatic segmentation of lesions using 2.5D U-net to achieve Dice coefficients of 0.74 ± 0.28 and 0.76 ± 0.29 with respect to manual delineation performed by two radiologists. The inter-observer variability measured by the Dice metric was 0.79 ± 0.25 between two radiologists. They calculated two imaging biomarkers, including the percentage of infection and average infectious HU for severity and progression assessment, resulting in AUC of 0.97. Thick-section CT imaging were recommended for high-pitch scans to decrease the acquisition time and motion artifacts (due to breath-holding) and reduce radiation doses to patients (10, 71). In our dataset, various slice thicknesses (1-8 mm) have been included to train a robust network against this parameter, which highly impacts image manifestation. The relative error of volume difference for percentage of infectious (lesion/lung) and relative mean HU Diff (%) were 0.22±6.3 % (95% CI: −0.95 - 1.4%) and −0.18 ± 3.4% (95% CI: −0.8 - %), demonstrating the high accuracy of COLI-Net for biomarker generation.

Potential foreseen applications are not limited to the detection and segmentation but could be useful in providing diagnostic and prognostic parameters calculated using lung and infections segmentation to estimate the percentage of infections, and enabling advanced image processing in COVID-19 patients. The existing body of research on pneumonia suggests that the pneumonia severity index (PSI) can potentially be used as a severity marker (72). A recent study classified COVID-19 patients into severe and non-severe patients based on PSI calculated using CT images (73). Different deep learning algorithms and radiomics analysis approaches using CT images have been examined recently for developing diagnostic (discriminating COVID-19 from bacterial/viral pneumonia) and prognostic (survival, hospital stay, intensive care unit (ICU) admission, risk of outcome) models, which require lung and lesion segmentation (18). Moreover, calculating the percentage of infection and well-aerated regions in the lung are frequently performed through visual assessment or by simply calculating HU values in the lungs, which is not only time-consuming but also lacks accuracy.

The established model exhibited noticeable performance variation across different COVID-19 patients collected from different countries, centers, with different patient backgrounds, and stages of the disease. Since the quality of CT images depends directly on the scanner model, imaging protocol (tube voltage, tube current, pitch factor, etc), and reconstruction algorithm, we employed various datasets from different centers to cover a large variability (10, 71). Though the proposed slgorithm was evaluated using a multi-center, multi-scanner, multi-national dataset and patients with a diverse background, stages of the disease, a full-scale adaptation of this model requires further clinical investigation and fine-tuning to the specific image acquisition parameters of a center. This framework provides multiple imaging biomarkers for COVID-19 patients to facilitate the assessment of their clinical relevance in diagnostic (discriminating COVID-19 from bacterial/viral pneumonia) and prognostic (survival, hospital stay, ICU admission, risk of outcome) applications. Further development should involve implementing lung lobes segmentation to calculate all potential imaging biomarkers at the lobes level.

## Conclusion

We set out to develop an automated algorithm capable of segmenting three-dimensional whole lung and infected regions in COVID-19 patients from chest CT images using deep learning techniques to enable fast, consistent, robust, and human error immune framework for lung and pneumonia lesion detection and delineation. Owing to the complex nature of the problem and high variability in lesion manifestation, transfer learning from whole lungs to pneumonia infection lesions was proposed and implemented to enrich specific COVID-19 pneumonia features identification from clinical studies. Moreover, a multi-centric and multi-scanner dataset was collected for the development of the deep learning model to establish an automated and generalizable platform for efficient COVID-19 patients management. The developed artificial intelligence model was evaluated using a wide range of COVID-19 patients of diverse populations with different stages of the disease from multiple centers around the world to enable big data analysis of COVID-19 for automated progression/regression assessment of pneumonia lesions in follow-up studies, provide diagnostic and prognostic metrics, and enable further advanced image processing.

## Data Availability

Part of data is available in mentioned link.

https://www.medseg.ai/

https://mosmed.ai/

http://medicalsegmentation.com/covid19/

## Acknowledgments

This work was supported by the Swiss National Science Foundation under grant SNRF 320030**_**176052.

## Supplemenatl Data

Number of true positive (TP), Number of false positive (FP), Number of false negative (FN), FN Number of true negative (FN), predicted segmentation (P), Manual segmentation (G)

**1. Dice Similarity Coefficient**

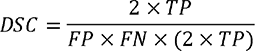

**2. Jaccard Index**

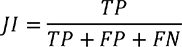

**3. False Negative**

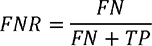

**4. False Positive**

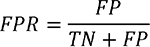

**5. Mean Surface Distance**

*d*(*p, G*) = distance between a point p belonging to the surface of a 3D surface predicted image (P) and its closest distance (MSD) between the two surfaces P and G can be defined as follows:

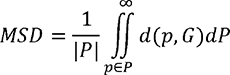

Where |P| denotes the surface area of P.

**6. Mean Harsdorf Distance**

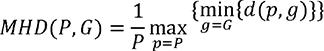

**7. Releive Difference Percentage**

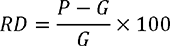

**8. Absolute difference Percentage**

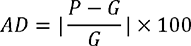

## Supplemental Figures

**Supplemental figure 1:**
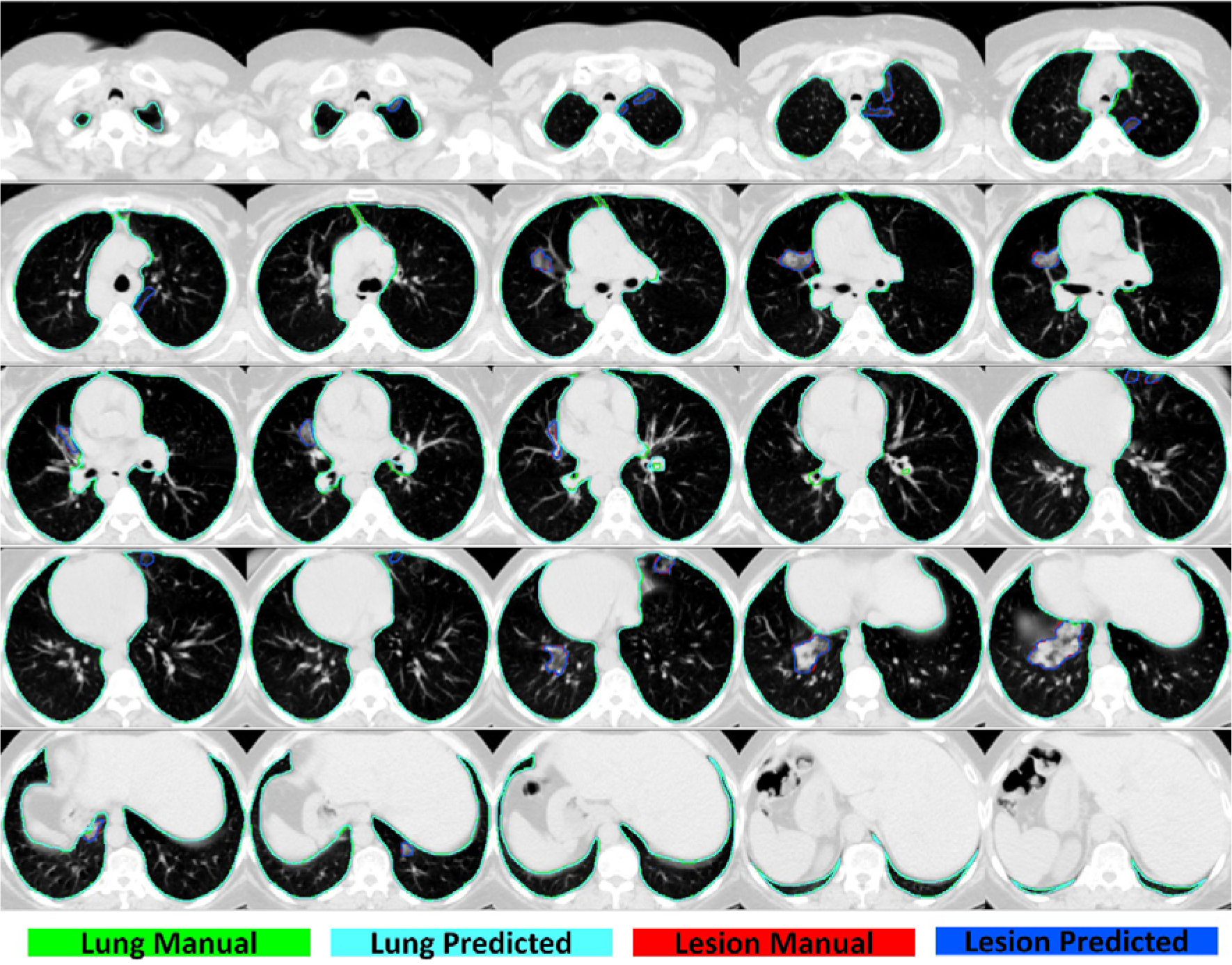
2D view of different slices of lung and lesion manual/predicted segmentation in Iran1 center, case 1.

**Supplemental figure 2:**
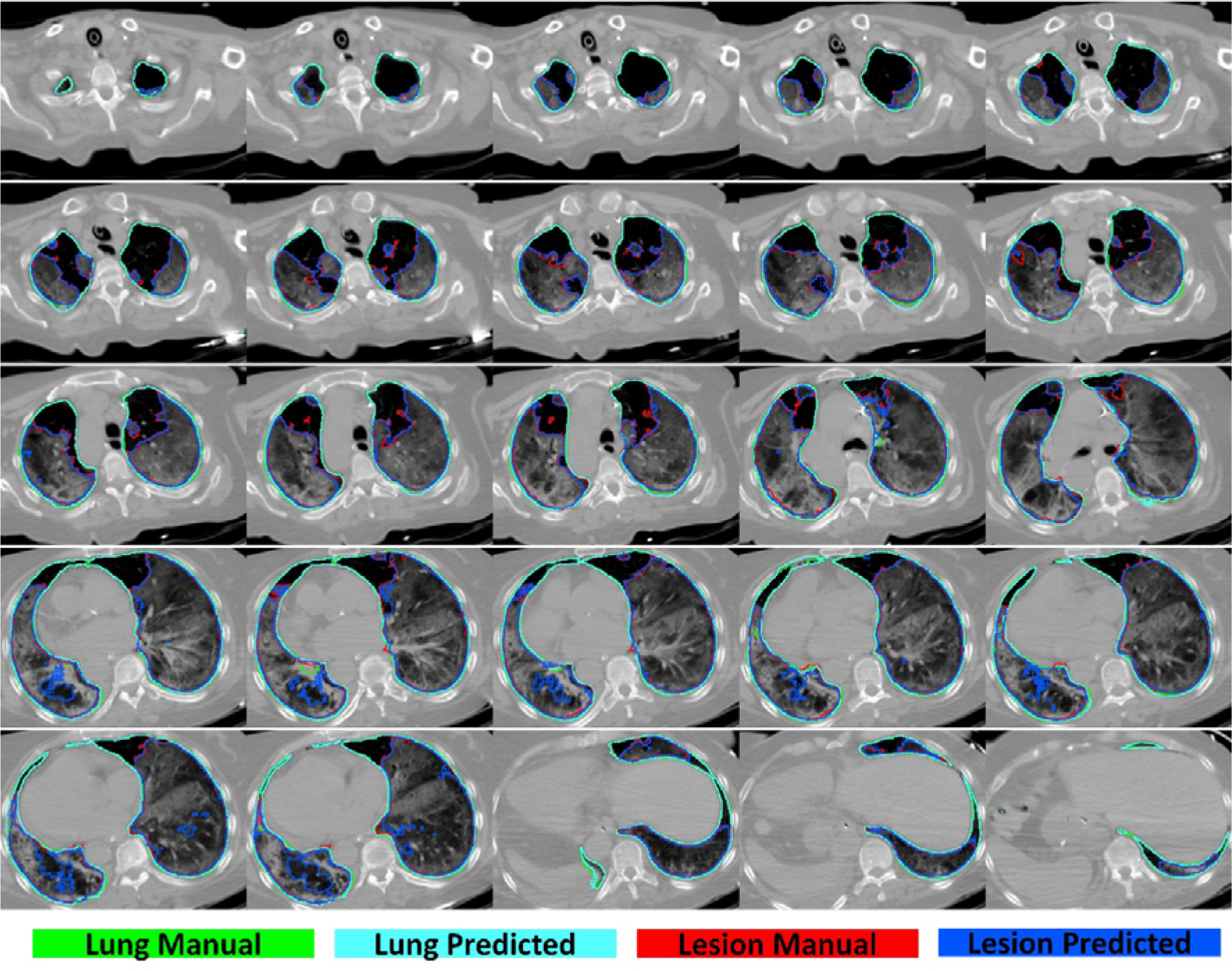
2D view of different slices of lung and lesion manual/predicted segmentation in Iran1 center, case 2.

**Supplemental figure 3:**
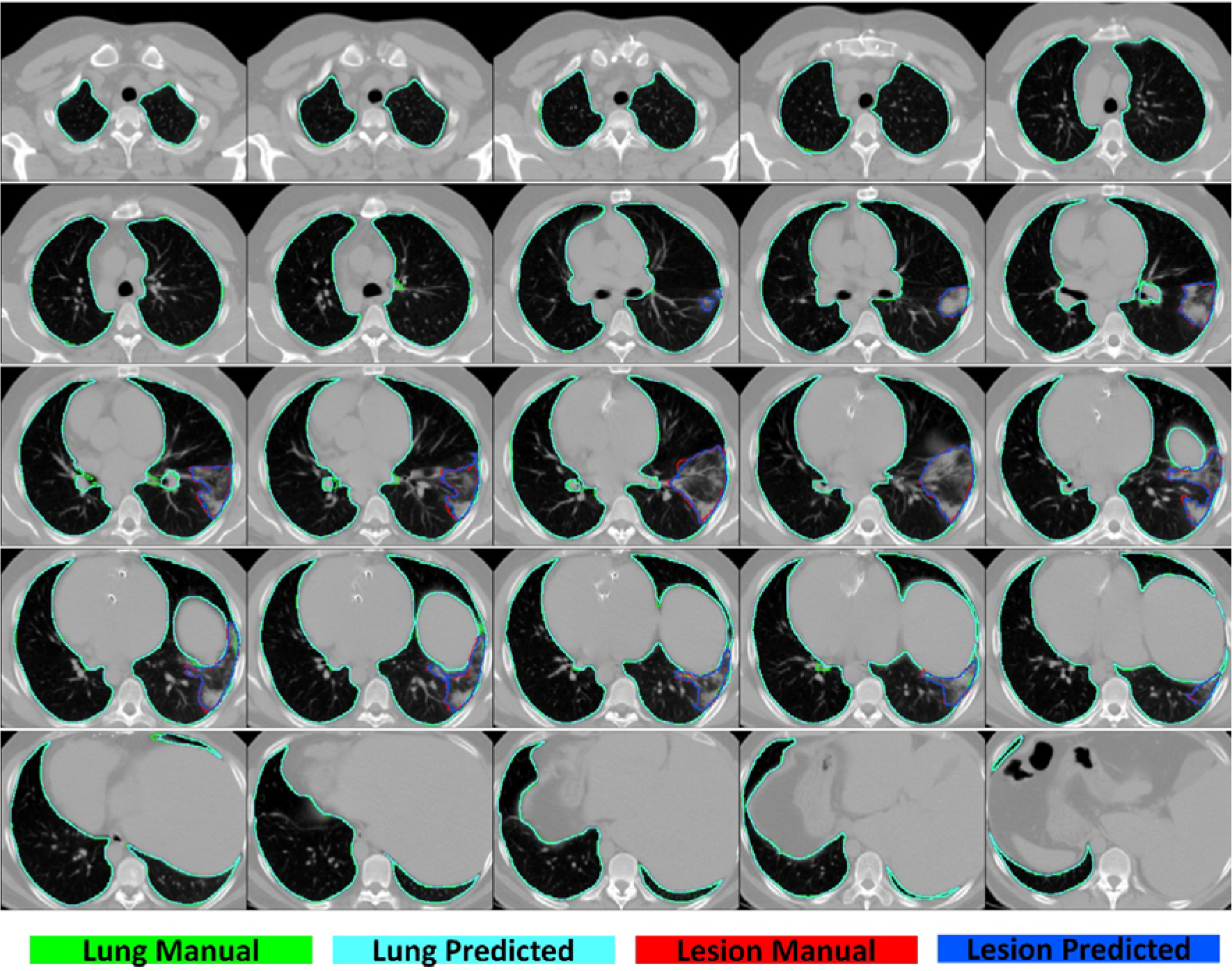
2D view of different slices of lung and lesion manual/predicted segmentation in Iran1 center, case 3.

**Supplemental figure 4:**
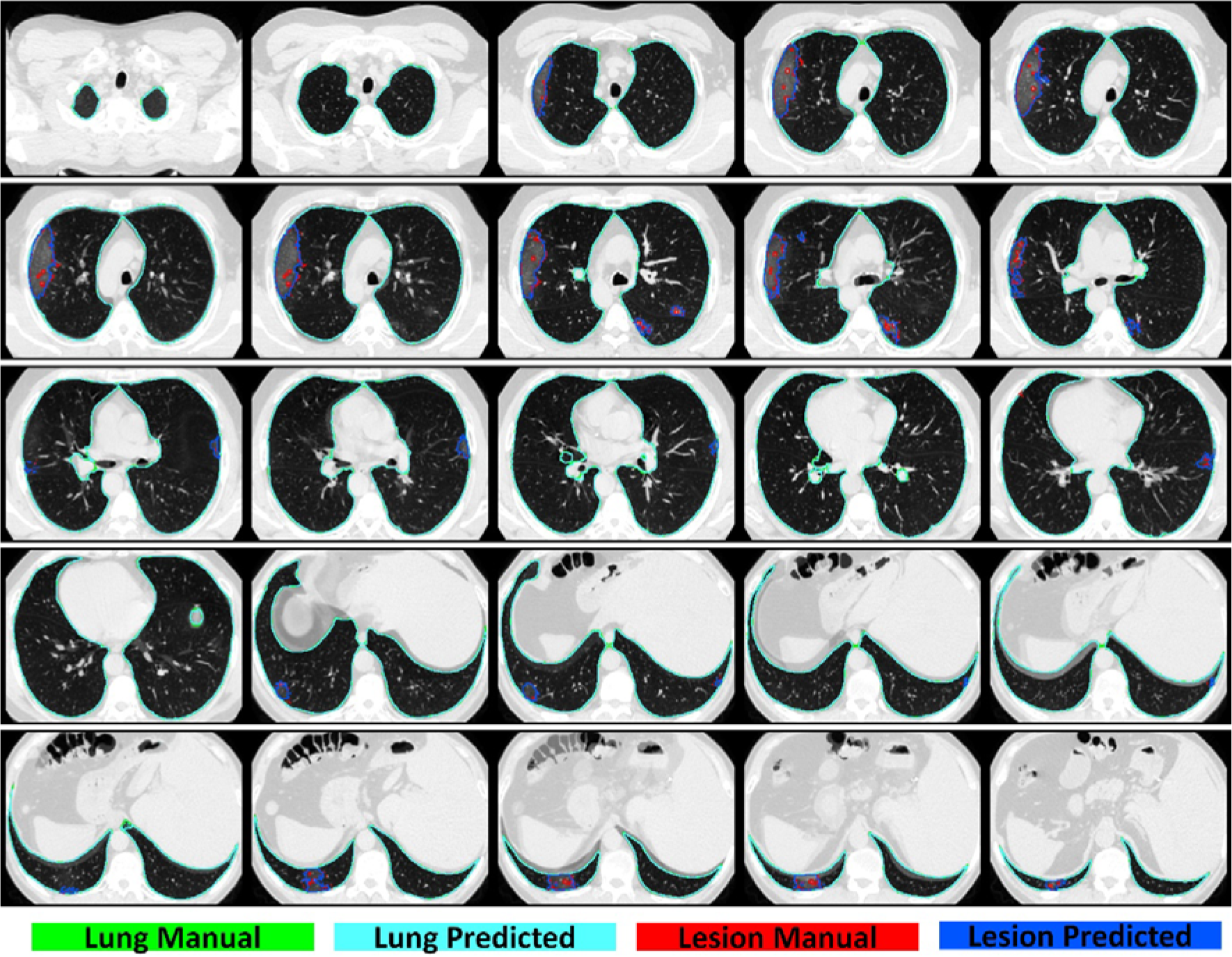
2D view of different slices of lung and lesion manual/predicted segmentation in Russia center, case 1.

**Supplemental figure 5:**
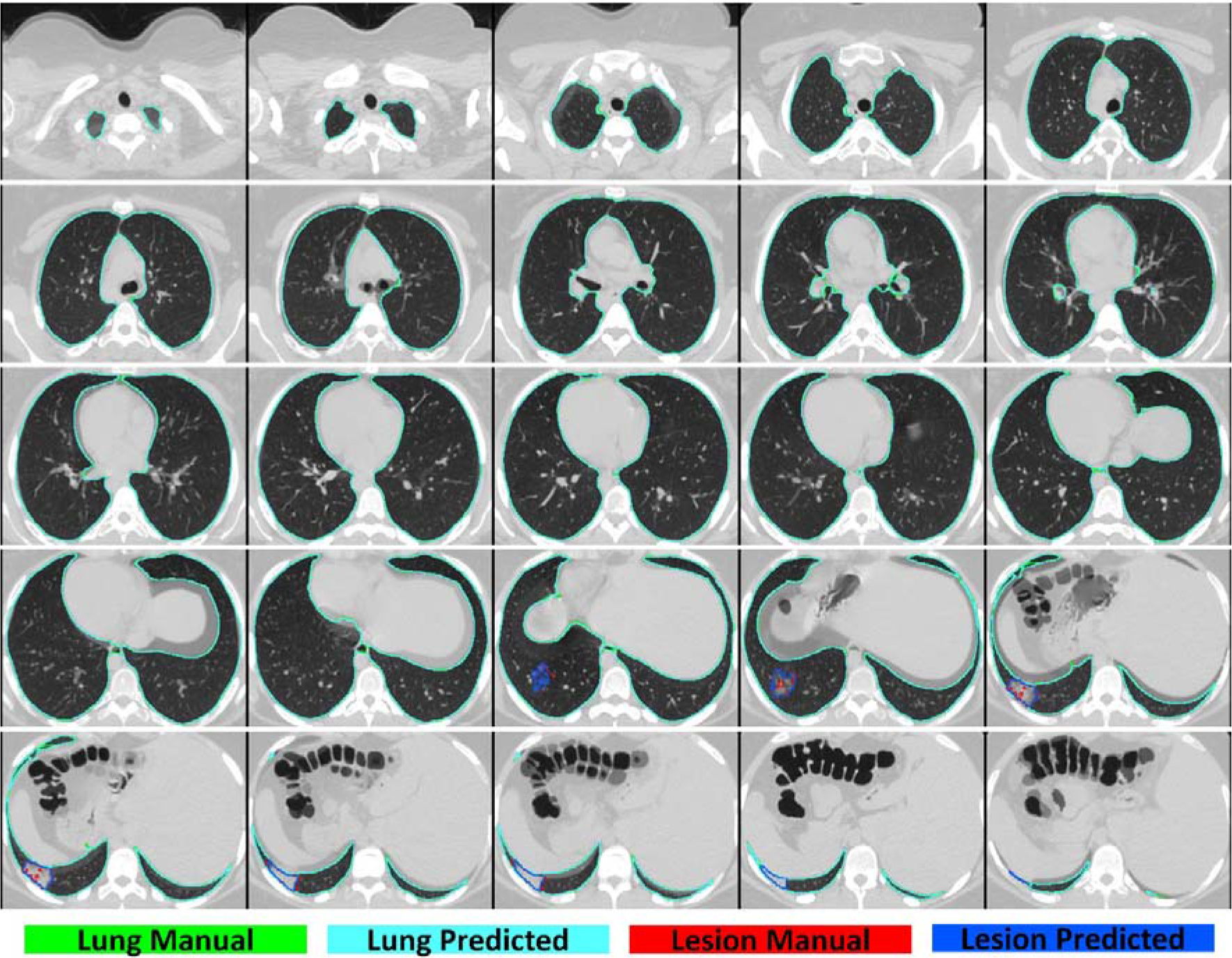
2D view of different slices of lung and lesion manual/predicted segmentation in Russia center, case 2.

**Supplemental figure 6:**
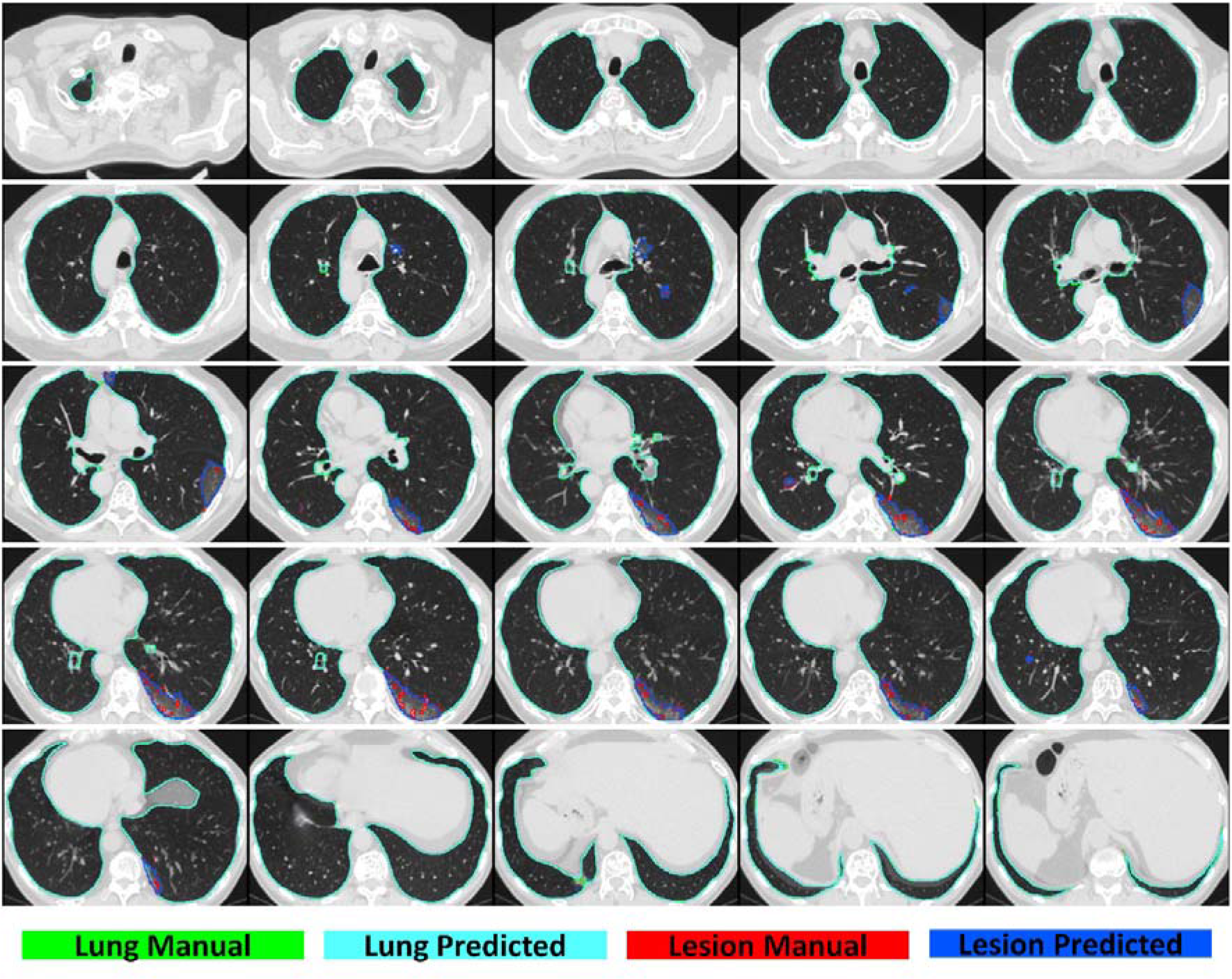
2D view of different slices of lung and lesion manual/predicted segmentation in Russia center, case 3.

**Supplemental figure 7:**
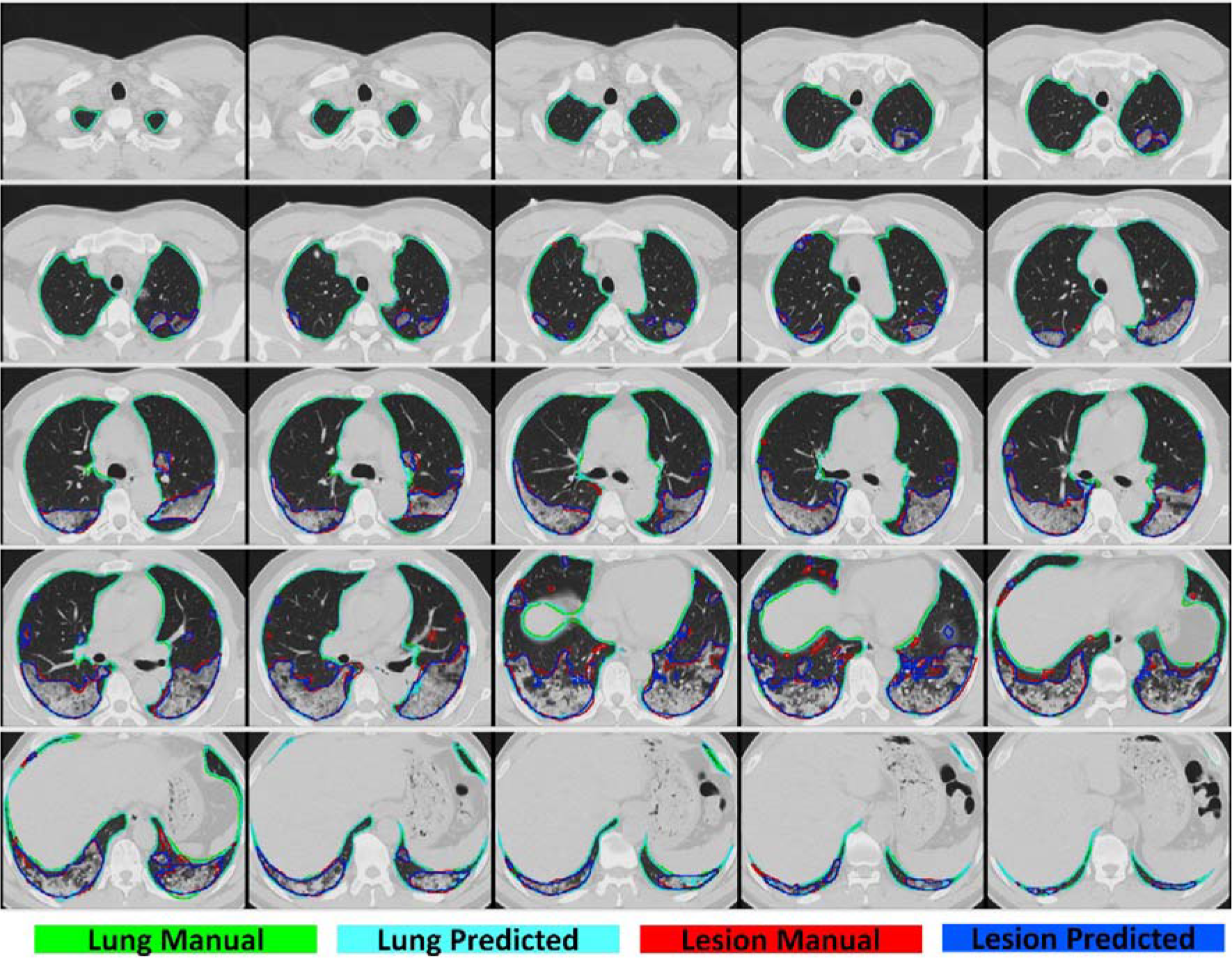
2D view of different slices of lung and lesion manual/predicted segmentation in Iran2 center, case 1.

**Supplemental figure 8:**
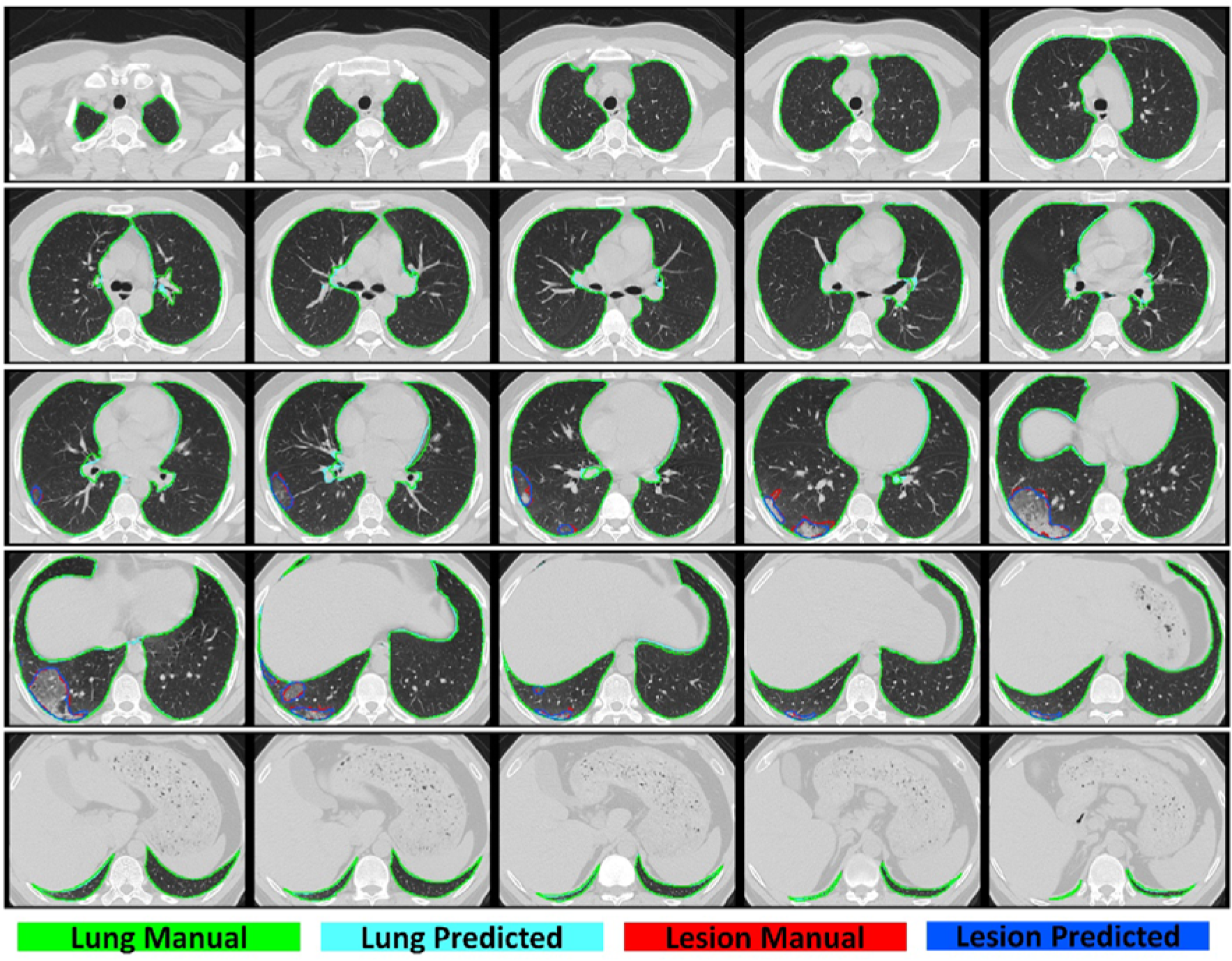
2D view of different slices of lung and lesion manual/predicted segmentation in Iran2 center, case 2.

**Supplemental figure 9:**
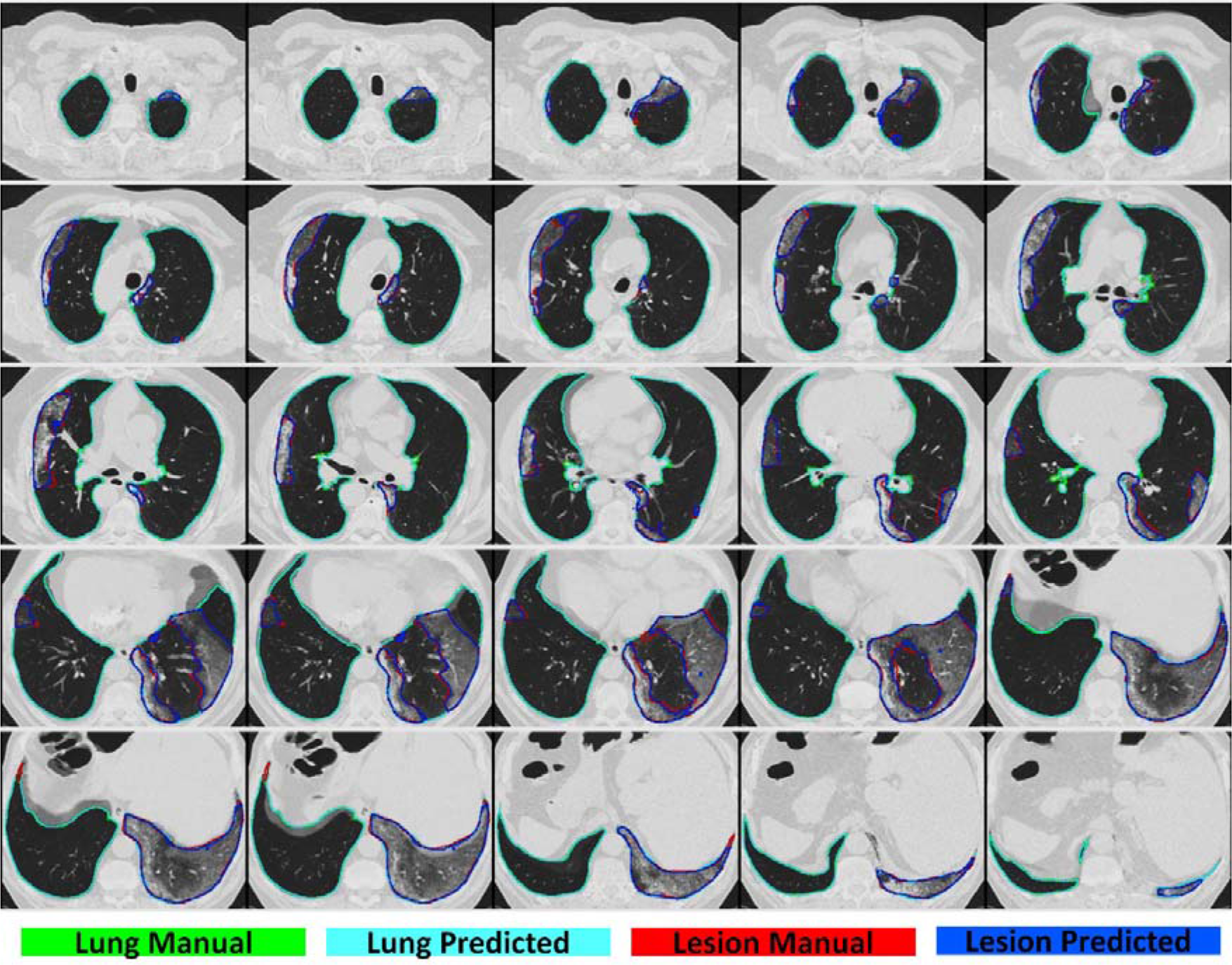
2D view of different slices of lung and lesion manual/predicted segmentation in China center, case 1.

**Supplemental figure 10:**
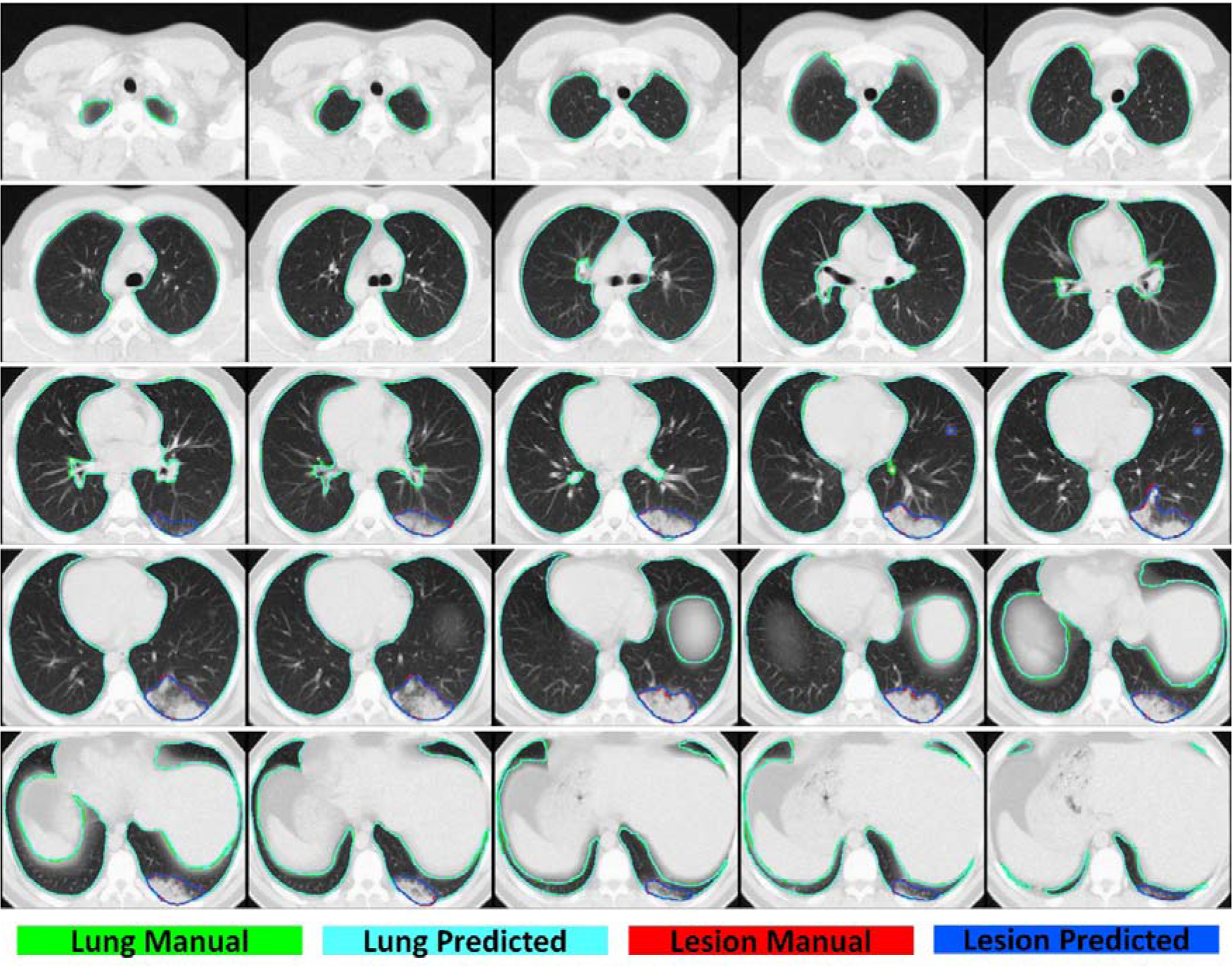
2D view of different slices of lung and lesion manual/predicted segmentation in China center, case 2.

**Supplemental figure 11:**
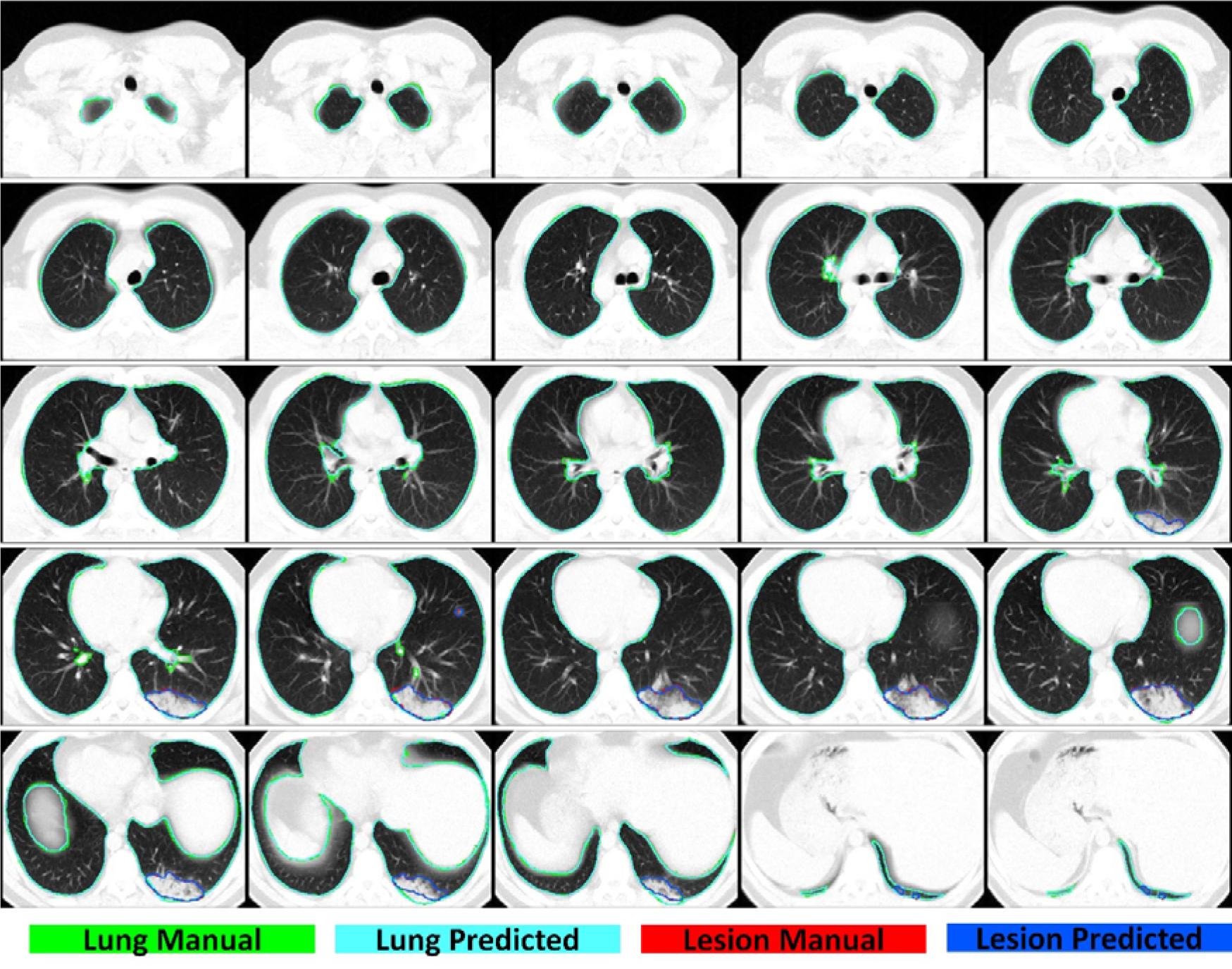
2D view of different slices of lung and lesion manual/predicted segmentation in Italy center, case 1.

**Supplemental figure 12:**
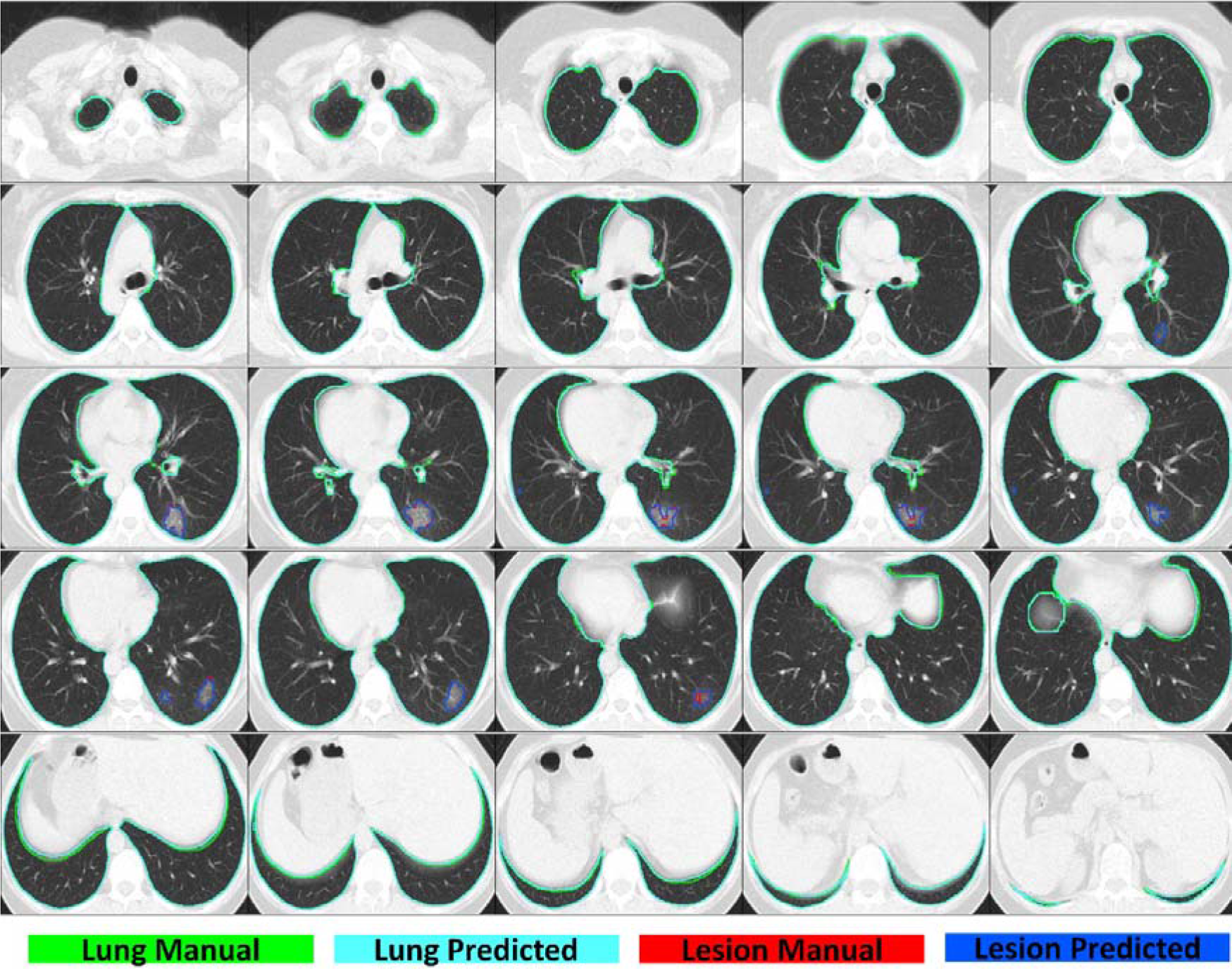
2D view of different slices of lung and lesion manual/predicted segmentation in Italy center, case 2.

**Supplemental figure 13:**
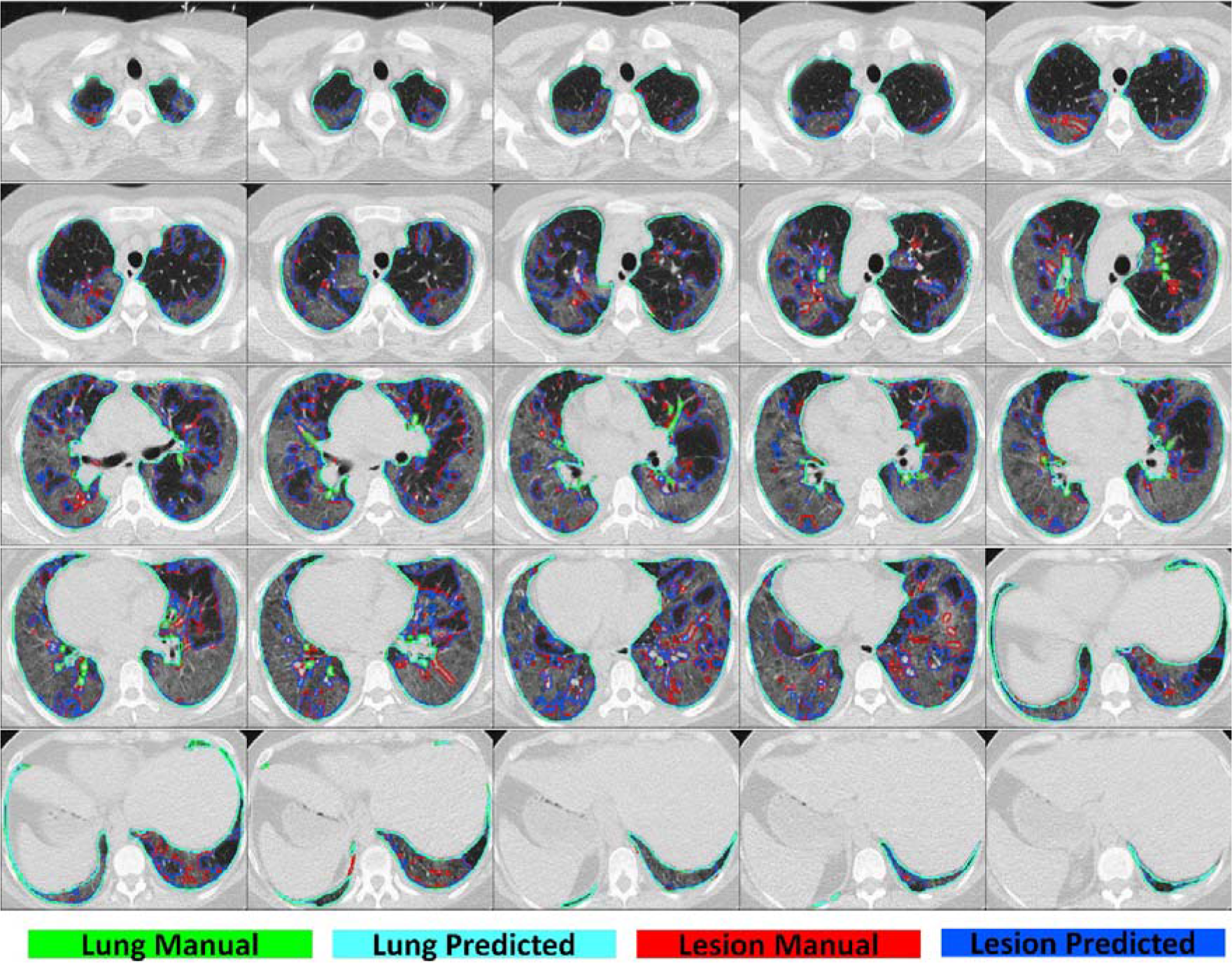
2D view of different slices of lung and lesion manual/predicted segmentation in Italy center, case 3.

**Supplemental figure 14:**
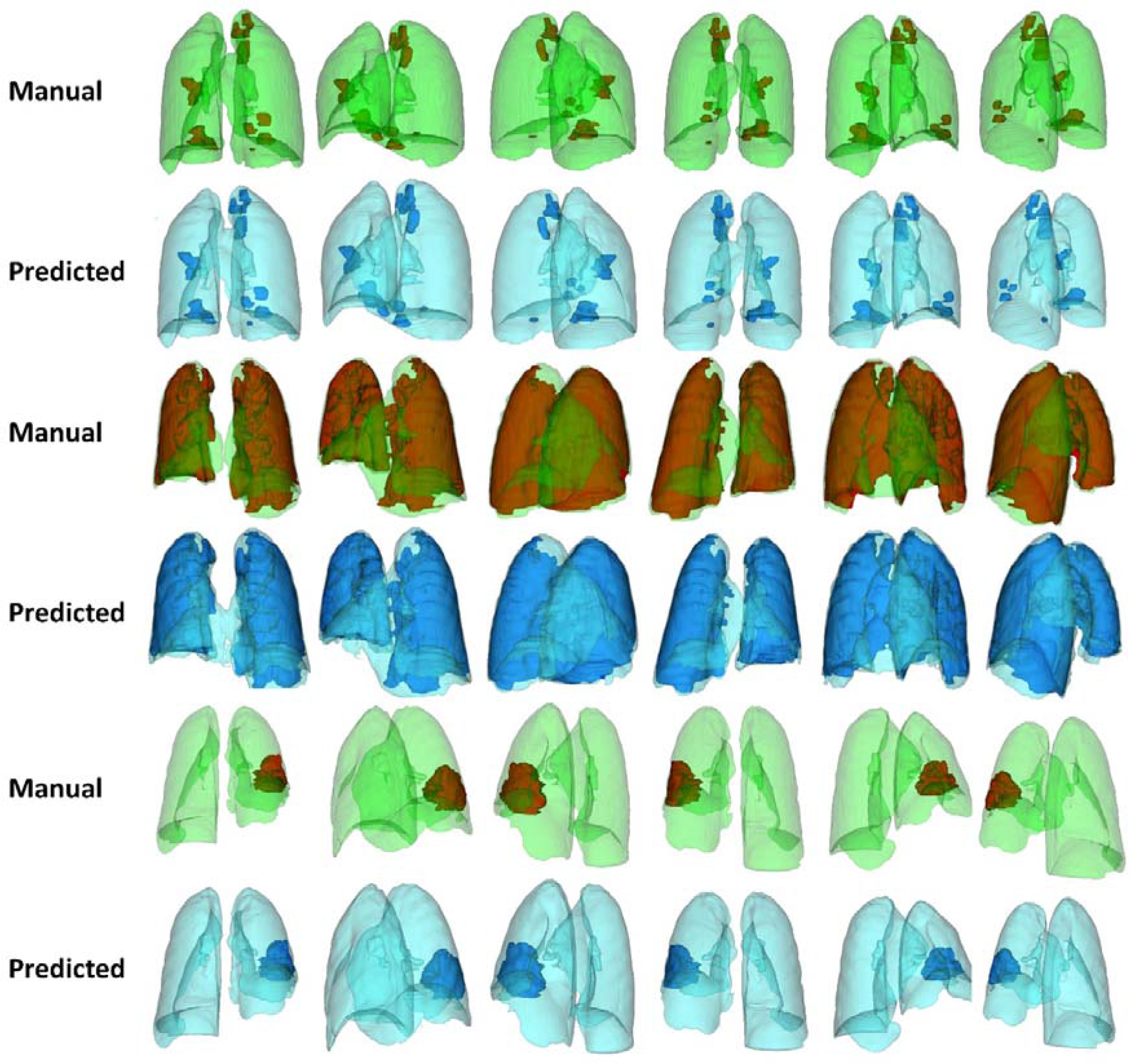
Manual and predicted segmentation 3D view for lung and lesion.

**Supplemental figure 15:**
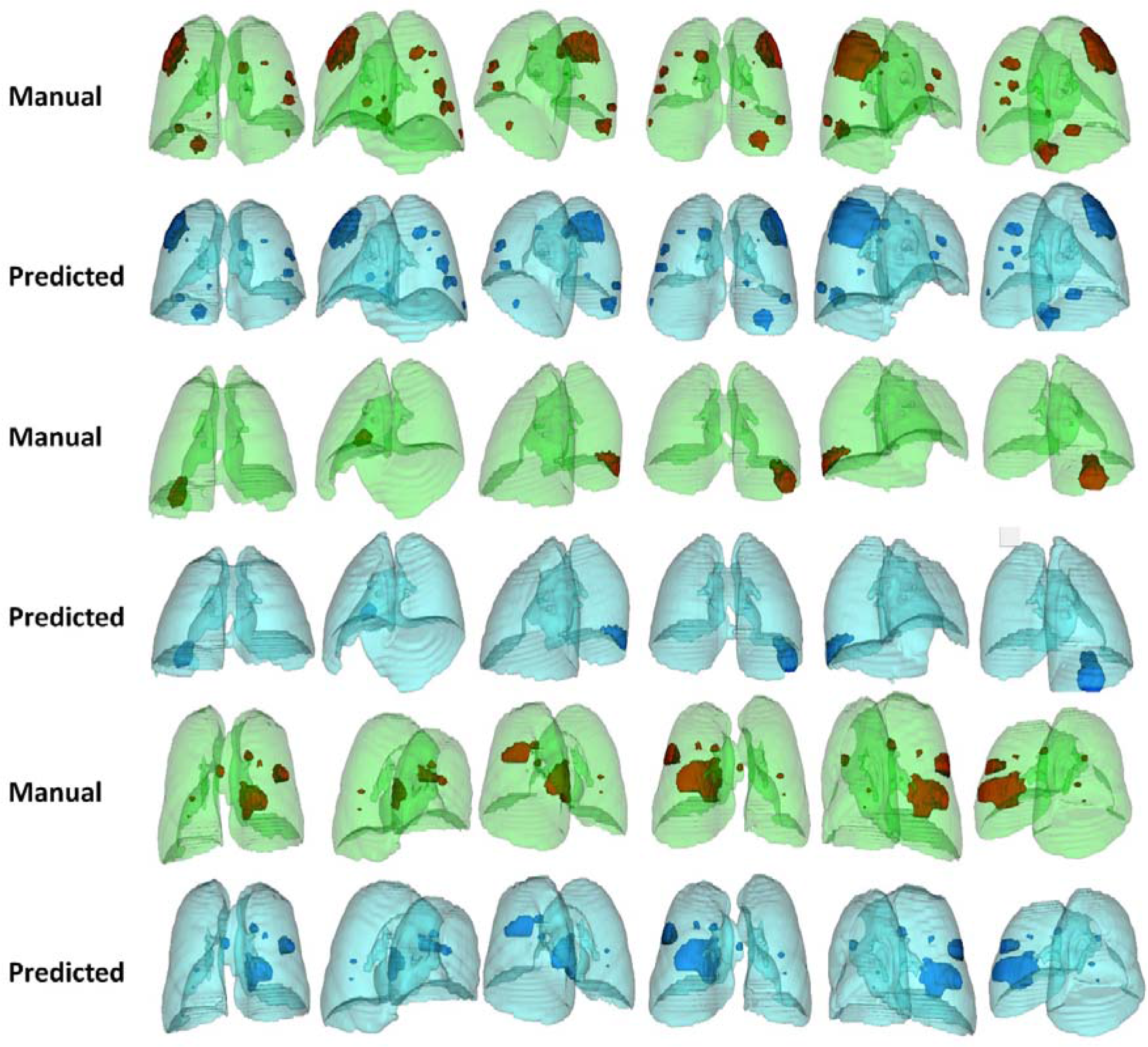
Manual and predicted segmentation 3D view for lung and lesion.

**Supplemental figure 16:**
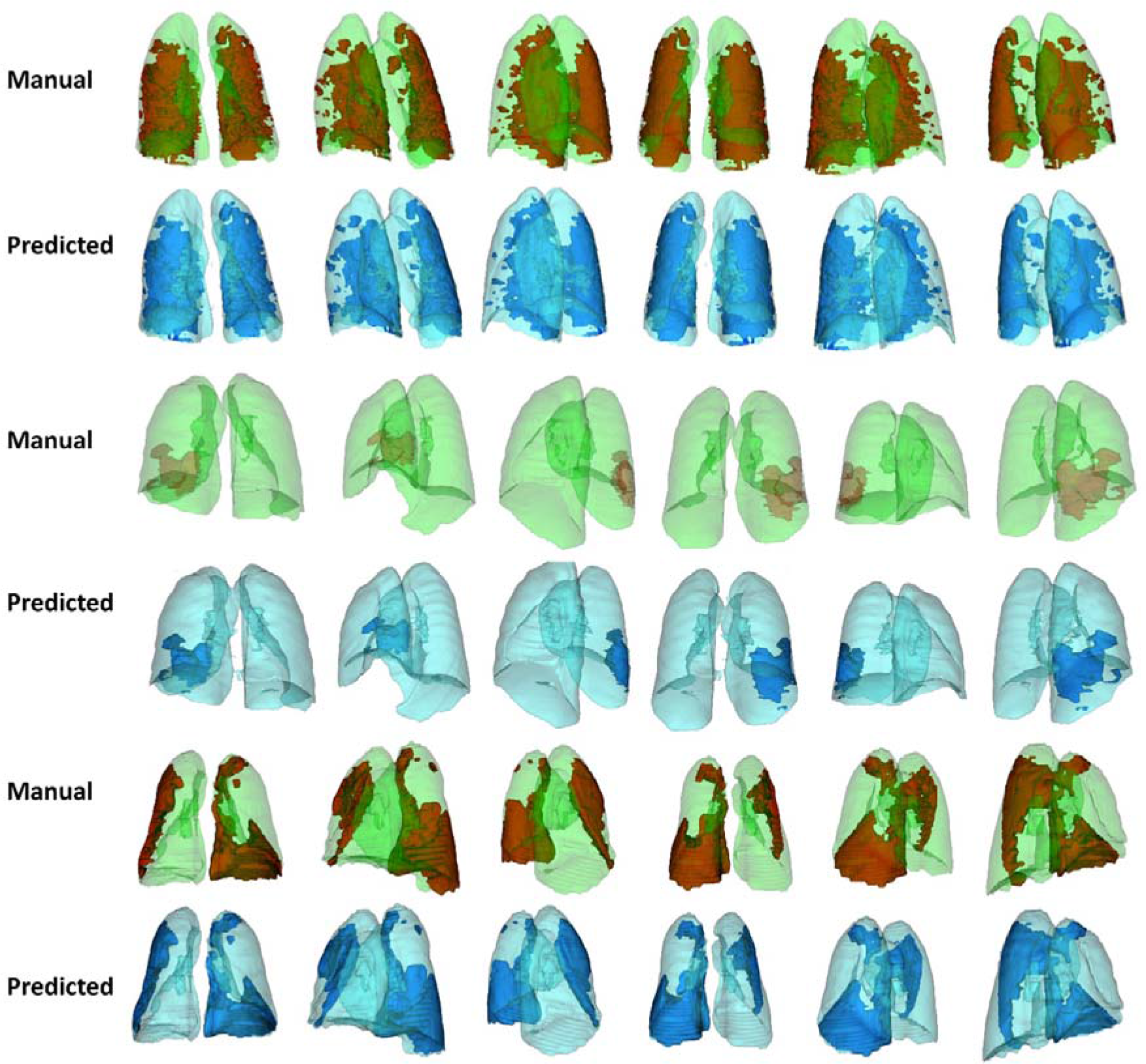
Manual and predicted segmentation 3D view for lung and lesion.

**Supplemental figure 17:**
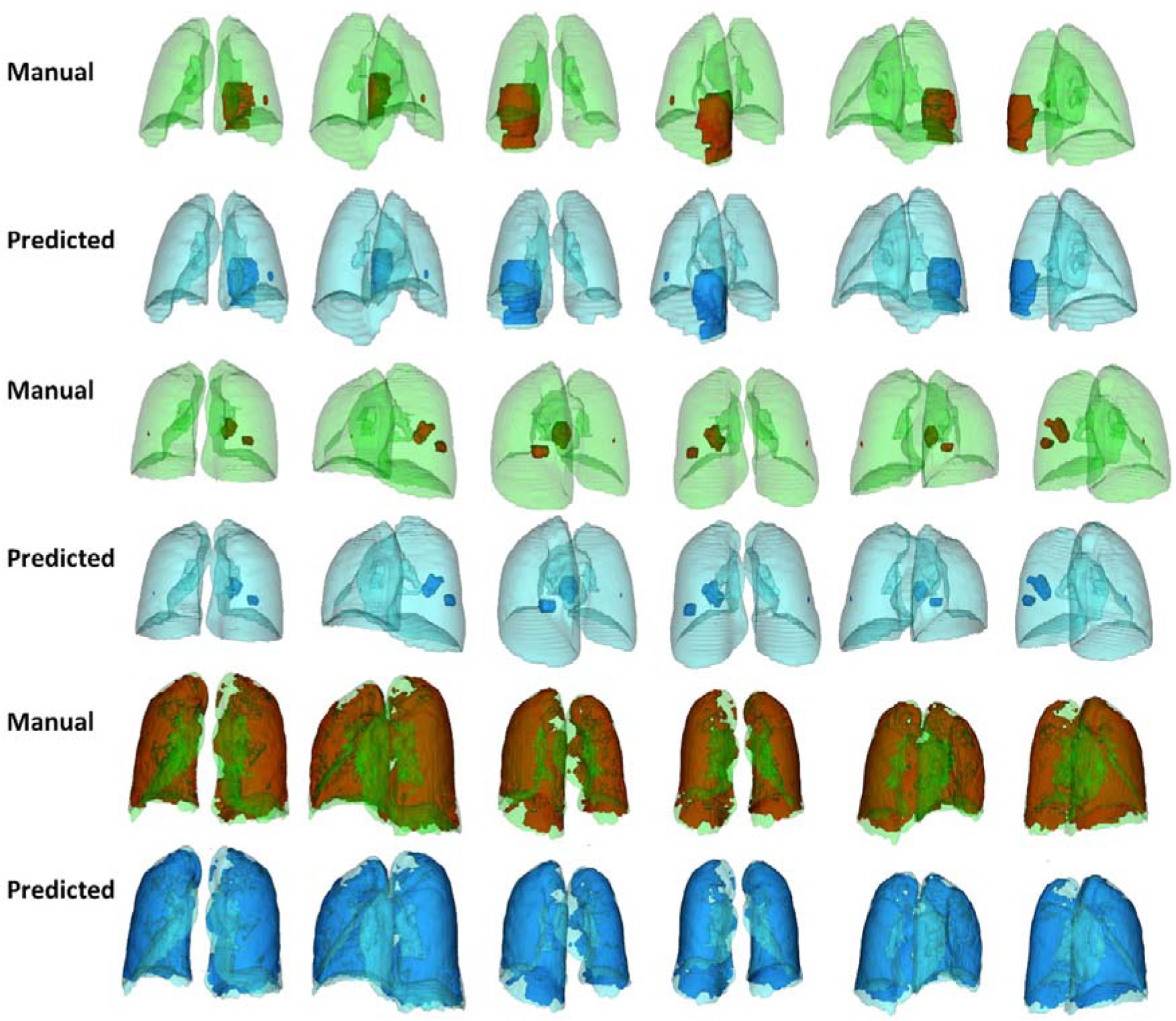
Manual and predicted segmentation 3D view for lung and lesion.

**Supplemental figure 18:**
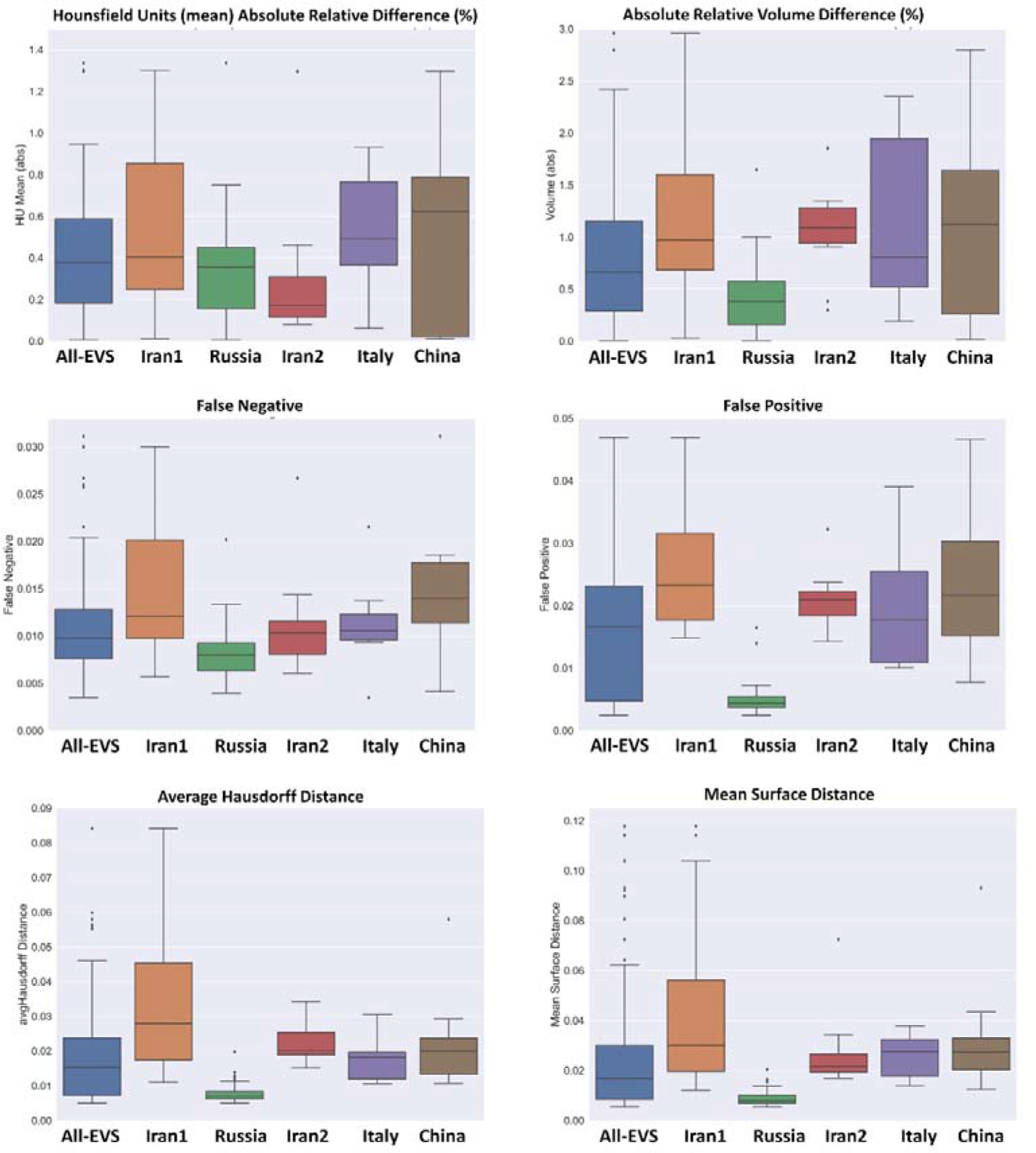
Box plots comparing various quantitative imaging metrics for lung segmentation, including Hounsfield unit(mean) absolute relative difference (%), absolute relative volume difference (%), false negative, false positive, average hausdorff distance and mean surface distance

**Supplemental figure 19:**
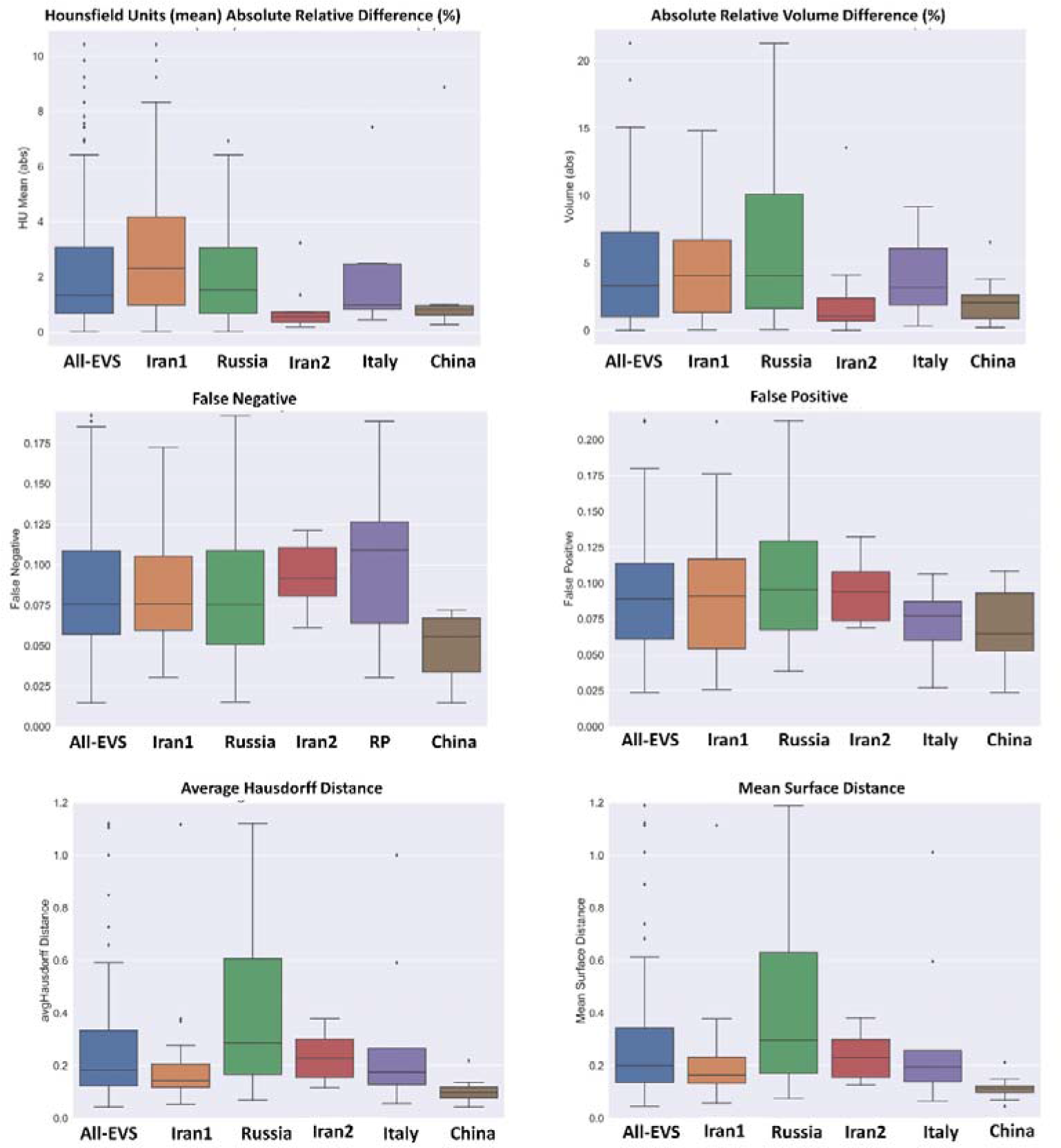
Box plots comparing various quantitative imaging metrics for infectious lesion segmentation, including Hounsfield unit(mean) absolute relative difference (%), absolute relative volume difference (%), false negative, false positive, average hausdorff distance and mean surface distance

**Supplemental figure 20:**
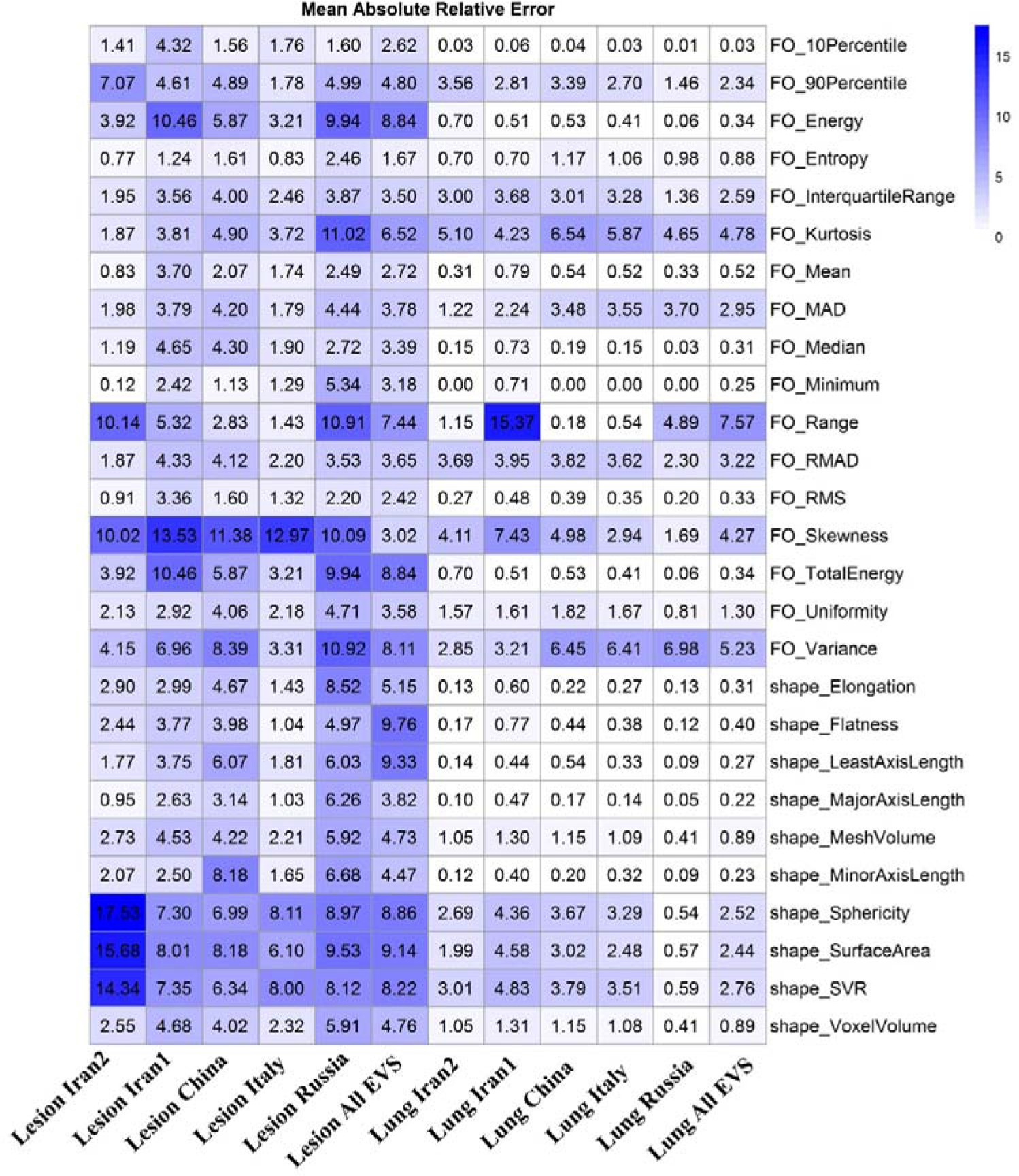
Figure 7: Mean absolute relative error of different first order (FO) and shape radiomics features for different datasets in lung and infection regions.

## Supplemental Tables

**Table 1:**
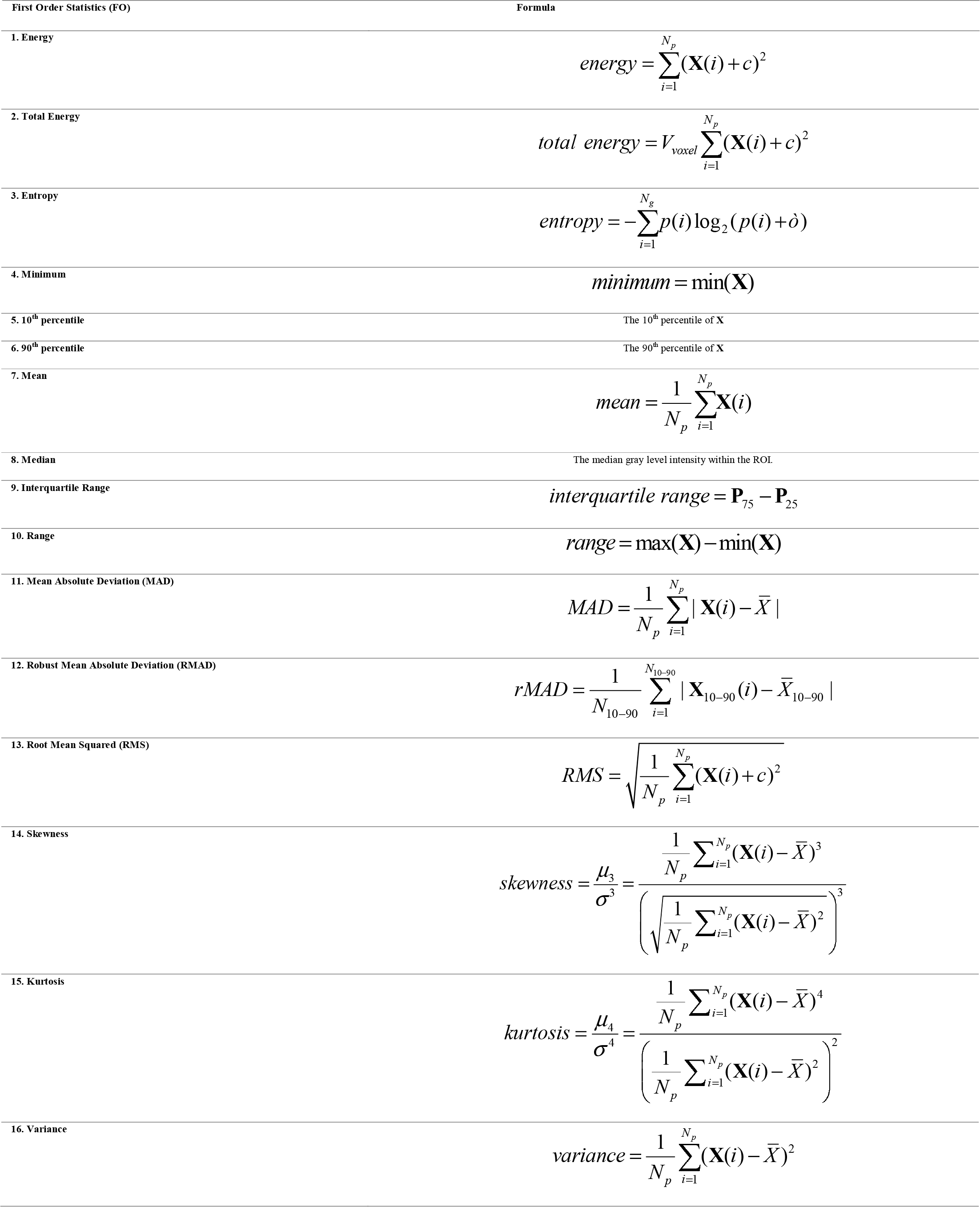
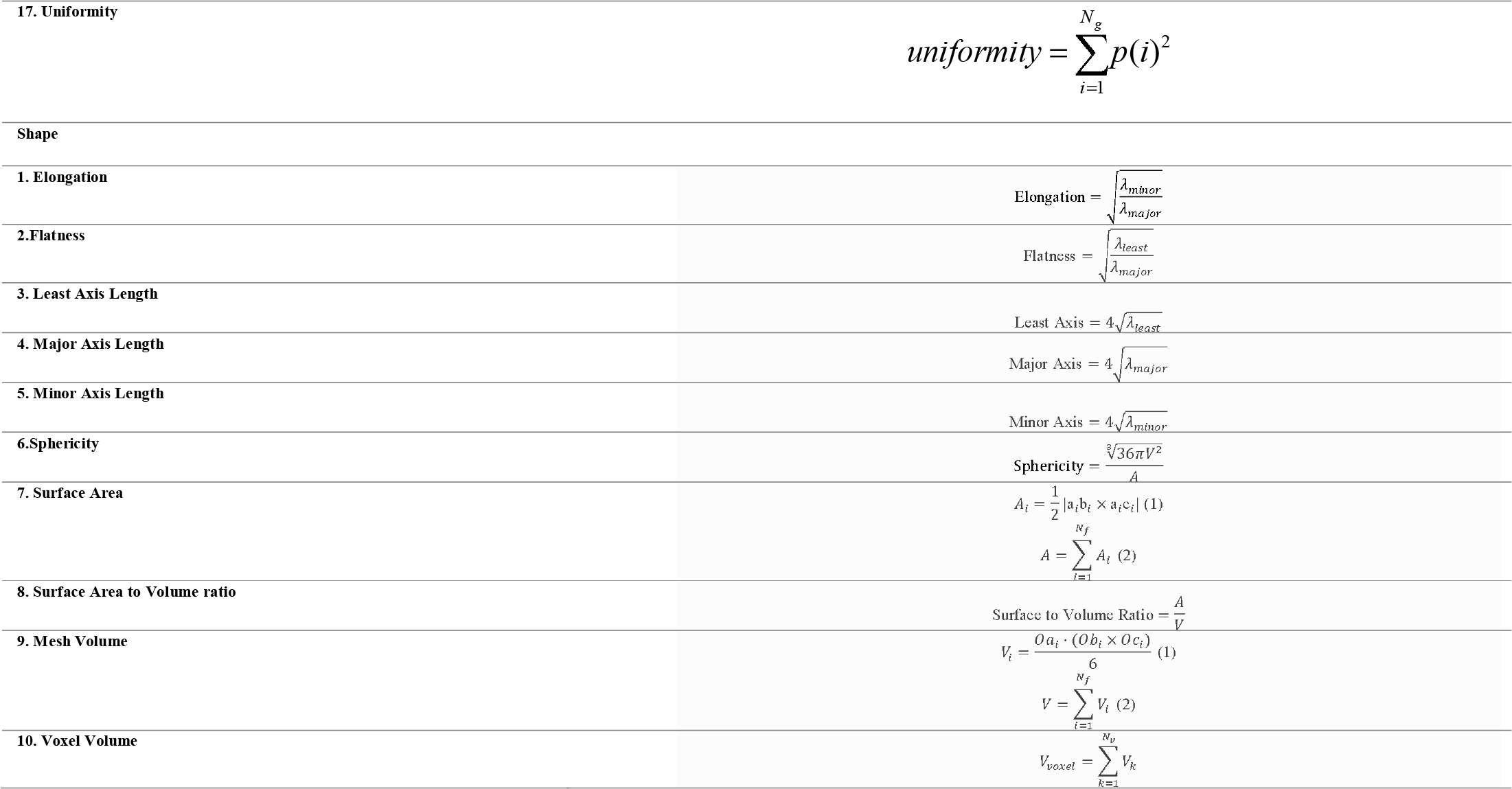
First order and shape radiomics features.

**Table 2:** Descriptive statistics of Dice Coefficient for lung and lesion in different datasets.

| **Dice Coefficient** | **Datasets** | **Min** | **Max** | **Mean ± SD** | **95% CI** |
| --- | --- | --- | --- | --- | --- |
| **Lung** | All EVS | 0.92 | 0.99 | $0.98 \pm 0.011$ | 0.98 - 0.99 |
| | China | 0.96 | 0.99 | $0.98 \pm 0.0086$ | 0.98 - 0.99 |
| | Italy | 0.97 | 0.99 | $0.98 \pm 0.0051$ | 0.98 - 0.99 |
| | Iran2 | 0.97 | 0.99 | $0.98 \pm 0.0046$ | 0.98 - 0.99 |
| | Russia | 0.99 | 0.99 | $0.99 \pm 0.0016$ | 0.99 - 0.99 |
| | Iran1 | 0.92 | 0.99 | $0.98 \pm 0.013$ | 0.97 - 0.98 |
| **Lesion** | All EVS | 0.8 | 0.98 | $0.91 \pm 0.038$ | 0.9 - 0.91 |
| | Italy | 0.92 | 0.98 | $0.94 \pm 0.024$ | 0.92 - 0.96 |
| | China | 0.85 | 0.97 | $0.91 \pm 0.04$ | 0.89 - 0.94 |
| | Iran2 | 0.83 | 0.93 | $0.90 \pm 0.031$ | 0.88 - 0.92 |
| | Russia | 0.8 | 0.96 | $0.90 \pm 0.044$ | 0.89 - 0.91 |
| | Iran1 | 0.84 | 0.97 | $0.91 \pm 0.03$ | 0.9 - 0.92 |

**Table 3:** Descriptive statistics of Jaccard Index for lung and lesion in different datasets.

| **Jaccard Index** | **Datasets** | **Min** | **Max** | **Mean ± SD** | **95% CI** |
| --- | --- | --- | --- | --- | --- |
| **Lung** | All EVS | 0.86 | 0.99 | $0.97 \pm 0.022$ | 0.97 - 0.97 |
| | China | 0.93 | 0.98 | $0.96 \pm 0.016$ | 0.95 - 0.97 |
| | Italy | 0.95 | 0.98 | $0.97 \pm 0.0098$ | 0.96 - 0.98 |
| | Iran2 | 0.95 | 0.98 | $0.97 \pm 0.0089$ | 0.96 - 0.97 |
| | Russia | 0.98 | 0.99 | $0.99 \pm 0.0032$ | 0.99 - 0.99 |
| | Iran1 | 0.86 | 0.98 | $0.95 \pm 0.025$ | 0.95 - 0.96 |
| **Lesion** | All EVS | 0.66 | 0.96 | $0.83 \pm 0.062$ | 0.82 - 0.84 |
| | Italy | 0.85 | 0.96 | $0.89 \pm 0.043$ | 0.86 - 0.92 |
| | China | 0.74 | 0.94 | $0.84 \pm 0.068$ | 0.8 - 0.89 |
| | Iran2 | 0.71 | 0.88 | $0.82 \pm 0.05$ | 0.79 - 0.85 |
| | Russia | 0.66 | 0.93 | $0.82 \pm 0.071$ | 0.8 - 0.84 |
| | Iran1 | 0.73 | 0.94 | $0.84 \pm 0.05$ | 0.82 - 0.85 |

**Table 4:** Descriptive statistics of False Negative for lung and lesion in different datasets.

| **False Negative** | **Datasets** | **Min** | **Max** | **Mean ± SD** | **95% CI** |
| --- | --- | --- | --- | --- | --- |
| **Lung** | All EVS | 0.0035 | 0.086 | $0.013 \pm 0.011$ | 0.011 - 0.015 |
| | China | 0.0041 | 0.031 | $0.015 \pm 0.0074$ | 0.0099 - 0.019 |
| | Italy | 0.0035 | 0.022 | $0.011 \pm 0.0048$ | 0.0083 - 0.015 |
| | Iran2 | 0.006 | 0.027 | $0.011 \pm 0.0059$ | 0.0077 - 0.015 |
| | Russia | 0.004 | 0.02 | $0.0082 \pm 0.0027$ | 0.0074 - 0.009 |
| | Iran1 | 0.0057 | 0.086 | $0.019 \pm 0.017$ | 0.014 - 0.024 |
| **Lesion** | All EVS | 0.015 | 0.23 | $0.086 \pm 0.044$ | 0.078 - 0.094 |
| | Italy | 0.015 | 0.072 | $0.05 \pm 0.022$ | 0.034 - 0.065 |
| | China | 0.03 | 0.19 | $0.1 \pm 0.054$ | 0.068 - 0.14 |
| | Iran2 | 0.061 | 0.23 | $0.11 \pm 0.047$ | 0.076 - 0.13 |
| | Russia | 0.015 | 0.21 | $0.087 \pm 0.05$ | 0.073 - 0.1 |
| | Iran1 | 0.031 | 0.17 | $0.084 \pm 0.034$ | 0.075 - 0.094 |

**Table 5:** Descriptive statistics of False Positive for lung and lesion in different datasets.

| **False Positive** |  | **Min** | **Max** | **Mean ± SD** | **95% CI** |
| --- | --- | --- | --- | --- | --- |
| **Lung** | All EVS | 0.0025 | 0.073 | $0.017 \pm 0.014$ | 0.014 - 0.019 |
| | China | 0.0077 | 0.047 | $0.023 \pm 0.012$ | 0.016 - 0.031 |
| | Italy | 0.01 | 0.039 | $0.019 \pm 0.0099$ | 0.013 - 0.026 |
| | Iran2 | 0.014 | 0.032 | $0.021 \pm 0.0048$ | 0.018 - 0.024 |
| | Russia | 0.0025 | 0.016 | $0.0049 \pm 0.0025$ | 0.0043 - 0.0056 |
| | Iran1 | 0.015 | 0.073 | $0.027 \pm 0.014$ | 0.023 - 0.032 |
| **Lesion** | All EVS | 0.024 | 0.32 | $0.098 \pm 0.055$ | 0.088 - 0.11 |
| | China | 0.024 | 0.11 | $0.069 \pm 0.029$ | 0.049 - 0.09 |
| | Italy | 0.027 | 0.11 | $0.07 \pm 0.029$ | 0.051 - 0.089 |
| | Iran2 | 0.069 | 0.13 | $0.093 \pm 0.022$ | 0.08 - 0.11 |
| | Russia | 0.039 | 0.32 | $0.11 \pm 0.068$ | 0.094 - 0.13 |
| | Iran1 | 0.026 | 0.24 | $0.094 \pm 0.048$ | 0.08 - 0.11 |

**Table 6:** Descriptive statistics of Average Hausdorff Distance for lung and lesion in different datasets.

| **Average Hausdorff Distance** | **Datasets** | **Min** | **Max** | **Mean ± SD** | **95% CI** |
| --- | --- | --- | --- | --- | --- |
| **Lung** | All EVS | 0.005 | 0.14 | $0.022 \pm 0.026$ | 0.018 - 0.027 |
| | China | 0.011 | 0.058 | $0.022 \pm 0.014$ | 0.014 - 0.031 |
| | Italy | 0.011 | 0.031 | $0.018 \pm 0.0062$ | 0.014 - 0.022 |
| | Iran2 | 0.015 | 0.034 | $0.022 \pm 0.0063$ | 0.019 - 0.026 |
| | Russia | 0.005 | 0.02 | $0.0077 \pm 0.0028$ | 0.0069 - 0.0085 |
| | Iran1 | 0.011 | 0.14 | $0.04 \pm 0.035$ | 0.03 - 0.051 |
| **Lesion** | All EVS | 0.043 | 5.6 | $0.42 \pm 0.73$ | 0.29 - 0.55 |
| | Italy | 0.043 | 0.22 | $0.11 \pm 0.055$ | 0.068 - 0.14 |
| | China | 0.057 | 1 | $0.3 \pm 0.31$ | 0.099 - 0.5 |
| | Iran2 | 0.12 | 0.38 | $0.23 \pm 0.092$ | 0.17 - 0.29 |
| | Russia | 0.068 | 5.6 | $0.65 \pm 0.95$ | 0.38 - 0.92 |
| | Iran1 | 0.054 | 3.6 | $0.3 \pm 0.6$ | 0.13 - 0.48 |

**Table 7:** Descriptive statistics of Mean Surface Distance for lung and lesion in different datasets.

| **Mean Surface Distance** | **Datasets** | **Min** | **Max** | **Mean ± SD** | **95% CI** |
| --- | --- | --- | --- | --- | --- |
| **Lung** | All EVS | 0.0056 | 0.17 | $0.026 \pm 0.028$ | 0.021 - 0.031 |
| | China | 0.012 | 0.093 | $0.032 \pm 0.023$ | 0.018 - 0.047 |
| | Italy | 0.014 | 0.038 | $0.026 \pm 0.0085$ | 0.021 - 0.032 |
| | Iran2 | 0.017 | 0.072 | $0.028 \pm 0.017$ | 0.017 - 0.038 |
| | Russia | 0.0056 | 0.02 | $0.009 \pm 0.0032$ | 0.0081 - 0.0099 |
| | Iran1 | 0.012 | 0.17 | $0.045 \pm 0.038$ | 0.034 - 0.056 |
| **Lesion** | All EVS | 0.046 | 6.1 | $0.45 \pm 0.79$ | 0.31 - 0.59 |
| | Italy | 0.046 | 0.21 | $0.12 \pm 0.05$ | 0.082 - 0.15 |
| | China | 0.066 | 1 | $0.31 \pm 0.3$ | 0.11 - 0.51 |
| | Iran2 | 0.13 | 0.38 | $0.24 \pm 0.091$ | 0.18 - 0.29 |
| | Russia | 0.075 | 6.1 | $0.69 \pm 1$ | 0.4 - 0.99 |
| | Iran1 | 0.057 | 3.9 | $0.32 \pm 0.64$ | 0.14 - 0.51 |

**Table 8:** Descriptive statistics of relative mean Hounsfield Unit difference (%) for lung and lesion in different datasets.

| Relative Mean HU Diff (%) | Datasets | Min | Max | Mean ± SD | 95% CI |
| --- | --- | --- | --- | --- | --- |
| **Lung** | All EVS | -4.2 | 3.9 | $0.03 \pm 0.84$ | -0.12 - 0.18 |
| | Iran1 | -4.2 | 3.9 | $-0.15 \pm 1.3$ | -0.52 - 0.23 |
| | Russia | -0.7 | 1.3 | $0.27 \pm 0.31$ | 0.19 - 0.36 |
| | Iran2 | -0.46 | 1.3 | $-0.02 \pm 0.49$ | -0.32 - 0.28 |
| | China | -0.93 | 0.77 | $-0.11 \pm 0.63$ | -0.5 - 0.28 |
| | Italy | -1.3 | 0.94 | $-0.25 \pm 0.67$ | -0.69 - 0.19 |
| **Lesion** | All EVS | -9.8 | 10 | -0.8 - 0.44 | -0.18 ± 3.4 |
| | Iran1 | -9.8 | 10 | -1.5 - 1.1 | -0.18 ± 4.4 |
| | Russia | -6.9 | 4.6 | -1.2 - 0.42 | -0.38 ± 2.7 |
| | Iran2 | -3.2 | 0.74 | -1.2 - 0.27 | -0.45 ± 1.2 |
| | China | -2.5 | 7.4 | -1.5 - 2.4 | 0.46 ± 3 |
| | Italy | -1 | 8.9 | -1.8 - 2.9 | 0.55 ± 3.4 |

**Table 9:** Descriptive statistics of absolute relative mean Hounsfield Unit difference (%) for lung and lesion in different datasets.

| Absolute Relative Mean HU Diff (%) | Datasets | Min | Max | Mean ± SD | 95% CI |
| --- | --- | --- | --- | --- | --- |
| **Lung** | All EVS | 0.006 | 4.2 | $0.52 \pm 0.66$ | 0.4 - 0.64 |
| | Iran1 | 0.012 | 4.2 | $0.79 \pm 0.98$ | 0.5 - 1.1 |
| | Russia | 0.006 | 1.3 | $0.33 \pm 0.24$ | 0.26 - 0.4 |
| | Iran2 | 0.082 | 1.3 | $0.31 \pm 0.37$ | 0.081 - 0.54 |
| | China | 0.062 | 0.93 | $0.54 \pm 0.3$ | 0.36 - 0.73 |
| | Italy | 0.012 | 1.3 | $0.52 \pm 0.47$ | 0.21 - 0.83 |
| **Lesion** | All EVS | 0.026 | 10 | $2.4 \pm 2.5$ | 1.9 - 2.8 |
| | Iran1 | 0.026 | 10 | $3.2 \pm 3$ | 2.3 - 4.1 |
| | Russia | 0.036 | 6.9 | $2 \pm 1.8$ | 1.5 - 2.6 |
| | Iran2 | 0.18 | 3.2 | $0.83 \pm 0.91$ | 0.26 - 1.4 |
| | China | 0.46 | 7.4 | $2.1 \pm 2.2$ | 0.66 - 3.5 |
| | Italy | 0.27 | 8.9 | $1.7 \pm 2.9$ | -0.27 - 3.7 |

**Table 10:** Descriptive statistics of relative volume difference (%) for lung and lesion in different datasets.

| Relative Volume Diff (%) | Datasets | Min | Max | Mean ± SD | 95% CI |
| --- | --- | --- | --- | --- | --- |
| **Lung** | All EVS | -3.1 | 6.4 | $0.38 \pm 1.2$ | 0.16 - 0.59 |
| | Iran1 | -3.1 | 6.4 | $0.83 \pm 1.5$ | 0.38 - 1.3 |
| | Russia | -1.6 | 0.86 | $-0.33 \pm 0.41$ | -0.44 - -0.21 |
| | Iran2 | -0.3 | 1.9 | $0.99 \pm 0.59$ | 0.63 - 1.4 |
| | China | -0.79 | 2.4 | $0.92 \pm 1.1$ | 0.25 - 1.6 |
| | Italy | -1.1 | 2.8 | $0.8 \pm 1.2$ | 0.0045 - 1.6 |
| **Lesion** | All EVS | -14 | 21 | $0.81 \pm 6.6$ | -0.39 - 2 |
| | Iran1 | -14 | 15 | $1 \pm 6.1$ | -0.82 - 2.8 |
| | Russia | -14 | 21 | $1.7 \pm 8$ | -0.58 - 4.1 |
| | Iran2 | -14 | 4.1 | $-1.3 \pm 4.7$ | -4.2 - 1.6 |
| | China | -9.2 | 1.9 | $-3.6 \pm 3.8$ | 6.1 - -1.1 |
| | Italy | -0.63 | 6.5 | $2.2 \pm 2.2$ | 0.62 - 3.7 |

**Table 11:** Descriptive statistics of absolute relative volume difference (%) for lung and lesion in different datasets.

| Absolute Relative Volume Diff (%) | Datasets | Min | Max | Mean ± SD | 95% CI |
| --- | --- | --- | --- | --- | --- |
| **Lung** | All EVS | 0.0047 | 6.4 | $0.89 \pm 0.88$ | 0.73 - 1 |
| | Iran1 | 0.031 | 6.4 | $1.3 \pm 1.1$ | 0.97 - 1.6 |
| | Russia | 0.0047 | 1.6 | $0.41 \pm 0.32$ | 0.32 - 0.5 |
| | Iran2 | 0.3 | 1.9 | $1.1 \pm 0.46$ | 0.77 - 1.3 |
| | China | 0.19 | 2.4 | $1.1 \pm 0.81$ | 0.65 - 1.6 |
| | Italy | 0.016 | 2.8 | $1.1 \pm 0.95$ | 0.46 - 1.7 |
| **Lesion** | All EVS | 0.018 | 21 | $4.8 \pm 4.6$ | 4 - 5.6 |
| | Iran1 | 0.049 | 15 | $4.7 \pm 3.9$ | 3.6 - 5.9 |
| | Russia | 0.087 | 21 | $6 \pm 5.5$ | 4.4 - 7.6 |
| | Iran2 | 0.018 | 14 | $2.6 \pm 4.1$ | 0.036 - 5.1 |
| | China | 0.33 | 9.2 | $4 \pm 3.3$ | 1.9 - 6.1 |
| | Italy | 0.23 | 6.5 | $2.3 \pm 2$ | 0.9 - 3.7 |

**Table 12:** Descriptive statistics of relative and absolute relative volume difference lesion/lung (%)

| **Volume Diff**<br>**lesion/Lesion (%)** | **Datasets** | **Min** | **Max** | **Mean ± SD** | **95% CI** |
| --- | --- | --- | --- | --- | --- |
| **RE** | All | -14 | 16 | $0.22 \pm 6.3$ | -0.95 - 1.4 |
| | Iran1 | -14 | 14 | $0.46 \pm 6.2$ | -1.4 - 2.3 |
| | Russia | -14 | 16 | $1.3 \pm 7.3$ | -0.89 - 3.5 |
| | Iran2 | -14 | 2.7 | $-2.3 \pm 4.6$ | -5.1 - 0.55 |
| | China | -9.6 | -0.0047 | $-4.4 \pm 3.3$ | -6.5 - -2.2 |
| | Italy | -0.65 | 6.3 | $1.5 \pm 2.3$ | -0.15 - 3.1 |
| **ARE** | All | 0.0047 | 16 | $4.7 \pm 4.2$ | 3.9 - 5.5 |
| | Iran1 | 0.23 | 14 | $5 \pm 3.7$ | 3.9 - 6.1 |
| | Russia | 0.23 | 16 | $5.5 \pm 4.8$ | 4.1 - 6.9 |
| | Iran2 | 0.034 | 14 | $2.8 \pm 4.2$ | 0.22 - 5.4 |
| | China | 0.0047 | 9.6 | $4.4 \pm 3.3$ | 2.2 - 6.5 |
| | Italy | 0.2 | 6.3 | $1.8 \pm 2$ | 0.39 - 3.2 |

**Table 13:** Descriptive statistics of relative volume (Lesion/Lung) for ground truth and predict images.

| **RV GT** | **Datasets** | **Min** | **Max** | **Mean ± SD** | **Mean (95% CI)** |
| --- | --- | --- | --- | --- | --- |
| **GT** | All | 0.00097 | 0.82 | $0.13 \pm 0.19$ | 0.095 - 0.16 |
| | Iran1 | 0.018 | 0.82 | $0.25 \pm 0.23$ | 0.18 - 0.32 |
| | Russia | 0.00097 | 0.088 | $0.018 \pm 0.016$ | 0.013 - 0.023 |
| | Iran2 | 0.014 | 0.3 | $0.085 \pm 0.091$ | 0.029 - 0.14 |
| | China | 0.0088 | 0.59 | $0.16 \pm 0.19$ | 0.04 - 0.28 |
| | Italy | 0.0064 | 0.56 | $0.12 \pm 0.19$ | -0.004 - 0.25 |
| **Predicted** | All | 0.0011 | 0.84 | $0.13 \pm 0.19$ | 0.094 - 0.16 |
| | Iran1 | 0.018 | 0.84 | $0.25 \pm 0.23$ | 0.18 - 0.32 |
| | Russia | 0.0011 | 0.081 | $0.018 \pm 0.015$ | 0.013 - 0.022 |
| | Iran2 | 0.014 | 0.29 | $0.082 \pm 0.088$ | 0.028 - 0.14 |
| | China | 0.0087 | 0.57 | $0.15 \pm 0.18$ | 0.037 - 0.27 |
| | Italy | 0.0064 | 0.56 | $0.13 \pm 0.19$ | -0.003 - 0.25 |

## Footnotes

1 http://medicalsegmentation.com/covid19/ https://www.medseg.ai/

